# A Functional Genomics Pipeline to Identify High-Value Asthma and Allergy CpGs in the Human Methylome

**DOI:** 10.1101/2022.05.19.22275204

**Authors:** Andréanne Morin, Emma E. Thompson, Britney A. Helling, Lyndsey E. Shorey-Kendrick, Pieter Faber, Tebeb Gebretsadik, Leonard B. Bacharier, Meyer Kattan, George T. O’Connor, Katherine Rivera-Spoljaric, Robert A. Wood, Kathleen C. Barnes, Rasika A. Mathias, Matthew C. Altman, Kasper Hansen, Cindy T. McEvoy, Eliot R. Spindel, Tina Hartert, Daniel J. Jackson, James E. Gern, Chris G. McKennan, Carole Ober, program collaborators for Environmental Influences on Child Health Outcomes and Children’s Respiratory and Environmental Workgroup

## Abstract

**Background:** DNA methylation of cytosines at CpG dinucleotides is a widespread epigenetic mark; but genome-wide variation has been relatively unexplored due to the limited representation of variable CpGs on commercial high-throughput arrays.

**Objective:** To explore this hidden portion of the epigenome, we combined whole-genome bisulfite sequencing (WGBS) with *in silico* evidence of gene regulatory regions to design a custom array of high-value CpGs. We focused these studies in airway epithelial cells from children with and without allergic asthma because these cells mediate the effects of inhaled microbes, pollution, and allergens on asthma and allergic disease risk.

**Methods:** We identified differentially methylated regions (DMRs) from WGBS in nasal epithelial cell (NEC) DNA from 39 children with and without allergic asthma of both European and African ancestries. We selected CpGs from DMRs, previous allergy or asthma Epigenome-Wide Association Studies (EWAS), or Genome-Wide Association Study (GWAS) loci, and overlapped them with functional annotations for inclusion on a custom Asthma&Allergy array. Using both the Custom and EPIC arrays, we performed EWAS of allergic sensitization (AS) in NEC DNA from children in the URECA birth cohort and using the Custom array in the INSPIRE birth cohort. We assigned each CpG on the arrays to its nearest gene and its promotor capture Hi-C interacting gene and performed expression quantitative trait methylation (eQTM) studies for both sets of genes.

**Results:** Custom array CpGs were enriched for intermediate methylation (IM) levels compared to EPIC CpGs. IM CpGs were further enriched among those associated with AS and for eQTMs on both arrays.

**Conclusions:** Our study revealed signature features of high-value CpGs and evidence for epigenetic regulation of genes at AS EWAS loci that are robust to race/ethnicity, ascertainment, age, and geography.

**Clinical Implications:** These studies identified allergic sensitization-associated differentially methylated CpGs and their target genes in airway epithelium, providing potential epigenetic mechanisms in the development of allergic diseases and suggesting novel drug targets.

**Capsule Summary:** This study of previously unexplored regions of the airway epithelial methylome revealed novel epigenetic mechanisms regulating genes previously implicated in the pathogenesis of asthma and allergic diseases.

## Introduction

Epigenetics refers to modifications of DNA molecules that do not alter the DNA sequence but play important roles in regulating gene expression. Environmental exposures can directly modify epigenetic marks in the human genome^1^, and epigenetic responses can mediate the effects of exposures on gene expression^2^ and disease risk^3^. Thus, the epigenome may contribute directly to disease risk or be sites of gene-environment interactions, providing both complementary data and mechanistic insights, respectively, to genome-wide association studies (GWAS). The most common epigenetic mark in the human genome is methylated cytosines at CpG dinucleotides and the availability of high-throughput array-based platforms to measure DNA methylation has led to an explosion of epigenome-wide association studies (EWAS)^4^. The most used commercial array, the InfinityMethylationEPIC BeadChip (Illumina, Inc., San Diego, CA), interrogates up to 850,000 CpGs, but this comprises <5% of CpGs in the human genome. Because the selection of CpGs on the EPIC array was agnostic with respect to disease or tissue types, findings to date may represent just the tip of the iceberg with an abundance of yet unknown functional CpG sites contributing to disease risk.

The goal of our study was to develop and validate a custom Asthma&Allergy DNA methylation array enriched for CpGs in airway epithelial cells that are likely to be functional and associated with asthma and allergic diseases in populations of diverse ancestries. We focused on airway epithelial cells because of their critical role in responding to inhaled microbes, pollution and allergens and mediating their downstream effects on asthma and allergic disease risk^5, 6^. An overview of our study design is shown in **Fig. 1**.

**Fig. 1.**
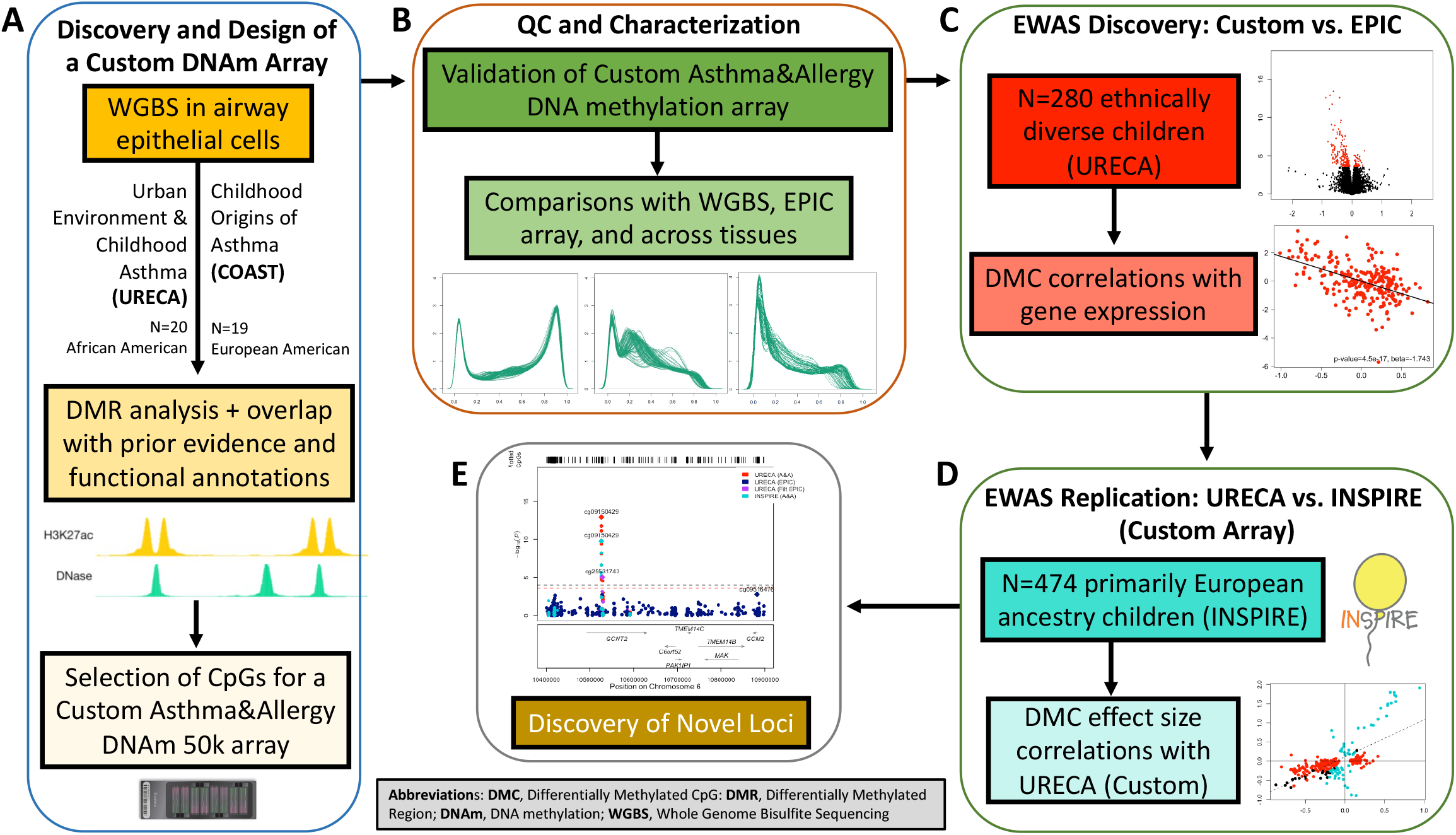
Overview of study design. **(A)** Whole-genome bisulfite sequencing (WGBS) and differential methylation were performed in airway epithelial cell DNA from birth cohorts comprised of African American children or European American young adults (half of each with allergic asthma and half without asthma or allergies). To select CpGs for the Custom array, we identified CpGs based on prior evidence of association with asthma or allergic disease from three sources (see text and **Fig. E1** in this article’s Online Repository). The CpGs were then prioritized based on their overlap with functional annotations to ultimately design a Custom array with 42,192 probes for 37,863 CpGs. **(B)** Airway epithelial cell DNA was hybridized to both the Custom array and the EPIC array, and β value distributions on the arrays were compared to the WGBS data and across tissues using the Custom and EPIC array. **(C-D)** We validated the Custom array by performing an EWAS of allergic sensitization (AS), examined correlations with the gene expression in the same cells, and replicated findings in a second cohort and with allergic asthma. **(E)** The combined results of the Custom and EPIC arrays in the discovery cohort and of the Custom array in the replication cohort for known and novel loci were examined in regional association plots.

## Methods

### Cohorts Included in Whole Genome Bisulfite Sequencing (WGBS) Studies or Epigenome-Wide Association Studies (EWAS)

Three longitudinal birth cohorts were included in these studies: the URban Environment and Childhood Asthma (URECA)^7^ cohort (n=20) and the Childhood Origins of ASThma (COAST)^8^ cohort (n=19) were included in the WGBS studies; URECA (n=280) and the Infant Susceptibility to Pulmonary Infections and Asthma Following RSV Exposure (INSPIRE)^9^ (n=474) were included in EWAS of allergic sensitization (AS). All studies were approved by the Institutional Review Boards of each of the institutions recruiting subjects. For each cohort, one parent or guardian provided informed consent for their child’s participation. Informed consent was obtained from participants in the COAST cohort after their 18^th^ birthday. The characteristics of the 280 URECA and 474 INSPIRE participants in the EWAS are described in **Table I**; detailed descriptions of the three cohorts are included in the Online Repository.

**Table I.**
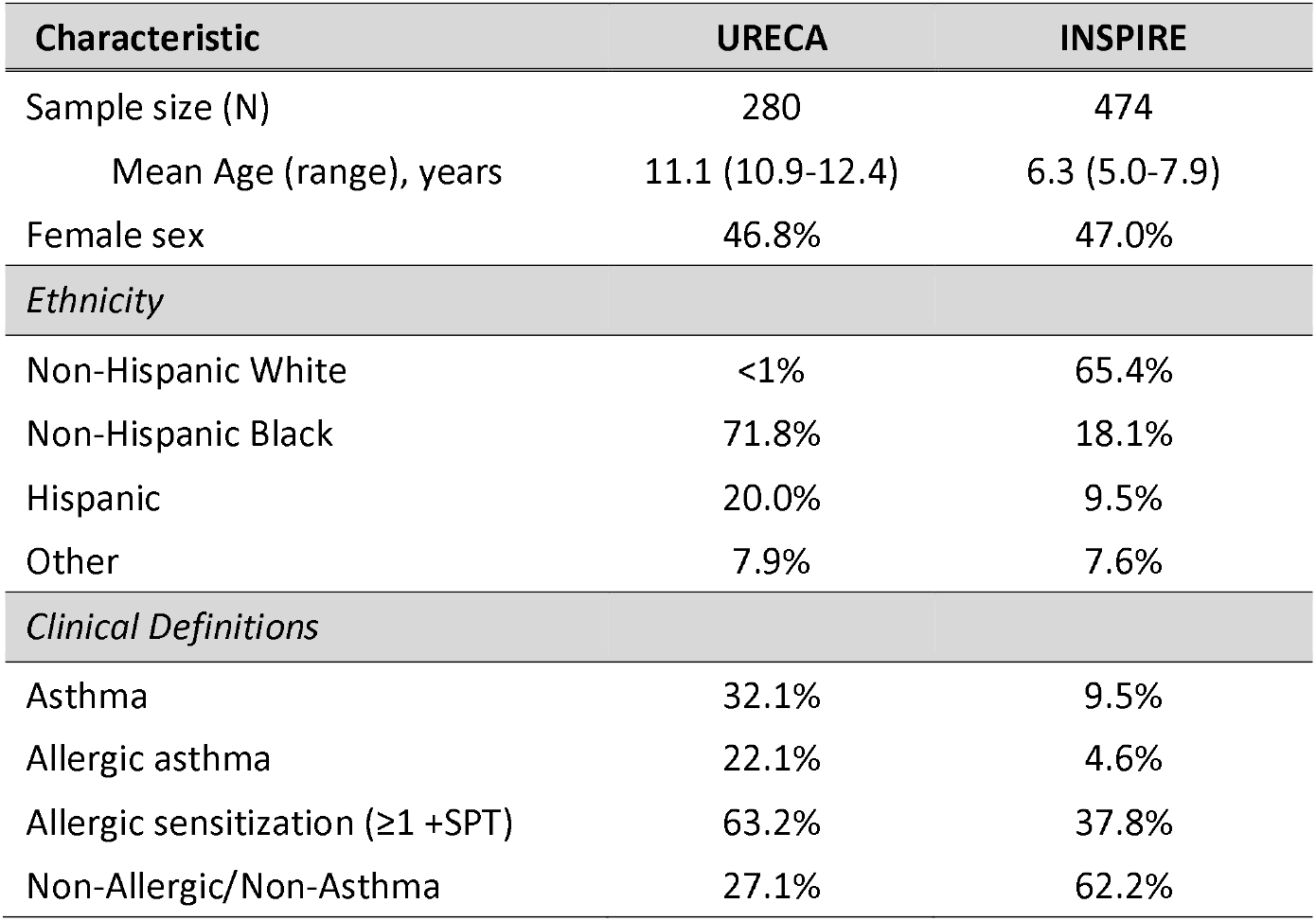
Demographic characteristics of URECA and INSPIRE samples. Also see Fig. E6 in this article’s Online Repository for details on allergic sensitizations.

| Characteristic | URECA | INSPIRE |
| --- | --- | --- |
| Sample size (N) | 280 | 474 |
| Mean Age (range), years | 11.1 (10.9-12.4) | 6.3 (5.0-7.9) |
| Female sex | 46.8% | 47.0% |
| *Ethnicity* |  |  |
| Non-Hispanic White | <1% | 65.4% |
| Non-Hispanic Black | 71.8% | 18.1% |
| Hispanic | 20.0% | 9.5% |
| Other | 7.9% | 7.6% |
| *Clinical Definitions* |  |  |
| Asthma | 32.1% | 9.5% |
| Allergic asthma | 22.1% | 4.6% |
| Allergic sensitization ($\geq 1$ +SPT) | 63.2% | 37.8% |
| Non-Allergic/Non-Asthma | 27.1% | 62.2% |

### Cohorts included in Cross-Tissue Comparison Studies

In addition to studying nasal epithelial cell DNA with the Asthma&Allergy Custom array in URECA, we included results from studies of the Custom array in DNA from nasal lavage cells (n=96), buccal cells (n=96), placenta (n=96) and cord blood (n=96). The nasal lavage cell DNA was isolated from URECA subjects at mean age 13.4 years (range = 12.8 - 14.9 years). The buccal, placenta and cord blood cell DNA was collected from participants in the Vitamin C to Decrease the Effects of Smoking in Pregnancy on Infant Lung Function (VCSIP) cohort^10^.

### Whole Genome Bisulfite Sequencing and DMR Studies

DNA (250 ng) from participants in the URECA and COAST cohorts was transferred to the University of Chicago Genomics Core Facility. Bisulfite conversion and sequencing were conducted using the EZ DNA Methylation-Gold Kit library prep and the Swift Biosciences Accel-NGS Methyl-seq DNA library kit, respectively. Samples were sequenced to a minimum of 330 million reads on the Illumina NovaSEQ6000 (S4 flowcell). Adapters were removed from sequence reads using trimgalore^11^ and then mapped using bismark (version 0.18.2)^12^ and the hg19 reference assembly. Bismark was also used to remove duplicate reads and to call methylation values. Prior to differential methylated region (DMR) analyses, CpGs were removed if they overlapped with a common single nucleotide polymorphism (SNP) (minor allele frequency [MAF] >0.05) in 1000 genomes CEU or YRI populations^13^ or in Blacklisted regions^14^. Variant calling from WGBS was performed using the biscuit algorithm, which also identified sample swaps and contamination. The variant calls from WGBS were compared to either array-based genotypes (COAST) or whole genome-sequence-based genotypes (URECA). All variant calls matched with an accuracy of >95%. One sample in COAST was a duplicate and was excluded from further analysis, leaving 19 samples for analysis (9 allergic asthmatics and 10 non-asthma/non-allergic controls). The DMR analyses are described in the Online Repository.

### Selection of CpGs and Design of the Custom Array

Our pipeline for selecting high-value CpGs for a custom array was implemented in three steps (**Fig. E1** in this article’s Online Repository). Additional details are described in the Online Repository. Our goal was to develop a custom array with 50,000 probes for CpGs that would complement and serve as a “booster” for the EPIC array in studies of asthma and allergic diseases. Briefly, we first identified a set of CpGs based on prior evidence of association with asthma or allergic diseases (atopic dermatitis/eczema, allergic rhinitis/hay fever, and food allergy) from three categories of studies. The first category included the 199,473 CpGs within DMRs from the WGBS study described above, the second included 19,057 CpGs associated with asthma or allergic disease in 17 previous EWAS (**Table E1** in this article’s Online Repository), and the third included 570,350 CpGs at 140 GWAS loci (**Table E2** in this article’s Online Repository). We then removed CpGs on the EPIC array, in ENCODE blacklisted regions^12^, and those that overlapped with a common SNP in 1000 Genomes European (CEU) or African (YRI) populations. In the second step, we further prioritize the remaining 696,225 CpGs, by considering their overlap with six functional annotations: 1) ENCODE^15^ TFBSs from all cell types; 2-4) ROADMAP Epigenetics^16^ transcriptional start sites, poised enhancers and active enhancers from smooth muscle (E078, E076, E103, E111), epithelial (E055, E056, E059, E061, E058), and blood cells (E062, E034, E045, E044, E043, E039, E041, E042, E040, E037, E048, E038, E047, E029, E050, E032, E046); 5) ATAC-seq in human cultured bronchial epithelial cells exposed to rhinovirus or a vehicle control from asthmatic and non-asthmatic individuals^17^, and 6) pcHi-C from ex vivo human bronchial epithelial cells^17^. We then required that CpGs within DMRs overlap with at least three annotations or other category of prior evidence, CpGs from prior EWAS overlap with at least one annotation or other category of prior evidence, and CpGs at GWAS loci overlap with at least four annotation categories or other category of prior evidence. After removing duplicates, 92,024 high-value CpGs remained and 53,700 of these CpGs were amenable to the Illumina design algorithm. We selected 44,047 CpGs targeted by 49,999 probes for manufacturing, of which 14% subsequently failed, resulting in a Custom array with 38,541 CpGs targeted by 43,605 probes. After removing probes with unknown origins and those without genomic coordinates, 37,863 CpGs passed array QC. The locations of the 37,863 CpGs, their selection among the three primary criteria, and their overlap with the annotation categories are shown in Supplementary Dataset 1. Finally, for our analyses we further removed probes that included a common SNP (MAF ≥5%) within 3 bp of the CpG interrogation site, probes that failed the minfi QC, and probes on the X chromosome, leaving 37,256 CpGs following both array and processing QC.

### Processing Custom and EPIC array methylation data in URECA and INSPIRE

DNA methylation was assessed using the Illumina Allergy&Asthma Custom BeadChip v1.0 or the Illumina Infinium MethylationEPIC BeadChip (Illumina, San Diego, CA) following bisulfite conversion at the University of Chicago Genomics Facility. Methylation data from both arrays were processed using minfi v1.29.1^18^. The annotation and manifest packages for the Allergy&Asthma array v1.0 can be found at https://github.com/hansenlab/IlluminaHumanMethylationAllergymanifest. The details of our protocols for processing the DNA methylation data on each array are described in the Online Repository.

### EWAS of AS in the URECA and INSPIRE cohorts

We defined AS as the percent of positive skin prick tests (14 airborne and oral allergens in URECA and 13 airborne allergens in INSPIRE). For each AS EWAS we used the following models:

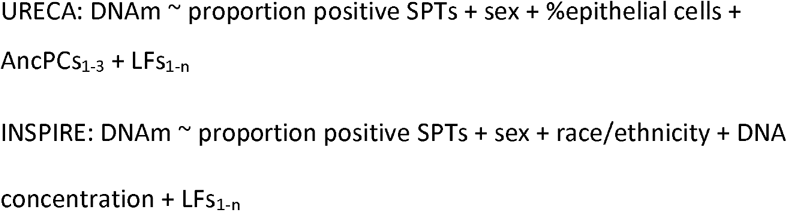

For the URECA EWAS, we included 14 and 5 latent variables for the Custom^19^ and EPIC^20^ arrays, respectively, after removing technical variables (see **Table E3** in this article’s Online Repository). Cell counts (300 cells/slide) were available on each sample^21^. Because we did not have information on cell proportion or genetic ancestry for INSPIRE, we included sex, parent-reported race/ethnicity, DNA concentration, and 21 latent factors as covariates to adjust for cell composition, as well as other unwanted variation in the EWAS. Analyses were performed in R (version 4.1.0) using limma v 3.50.0^22, 23^. To control the false discovery rate, we used a q-value threshold of 0.05. To assess the overlap of AS DMCs with allergic asthma DMCs, we tested the AS DMCs for association with allergic asthma in URECA and INSPIRE. For these studies, we defined allergic asthma cases as children with a diagnosis of asthma at age 10 (URECA) or age 6 (INSPIRE) <u>and</u> sensitization to at least one allergen (N=62 and 22, respectively). For these analyses, controls were defined as children without a diagnosis of asthma (ever) and without any positive skin prick tests (n=76 and 271, respectively). We calculated q-values based on the 193 AS DMCs in URECA and the 85 AS DMCs in INSPIRE.

### RNA-sequencing and expression Quantitative Trait Methylation (eQTM) studies in URECA

Protocols for processing samples for RNA-seq in nasal epithelial cells from the URECA children have been described^21^. Gene expression data were available in 249 of the children with DNA methylation data. In these samples, 15,643 genes were detected as expressed. To test for cis associations between DNA methylation and gene expression (eQTM studies), we used a linear model that included sex, percent epithelial cells, and three ancestry PCs as covariates (FDR <0.05).

## Results

### Identifying DMRs in Whole-Genome Bisulfite Sequences

We performed WGBS in airway epithelial cell DNA from 20 African American children (10 with allergic asthma, 10 without asthma or allergies; 11 years old) from the URECA cohort^7^ and 19 European American young adults (9 with allergic asthma, 10 without asthma or allergies; 18-20 years old) from the COAST cohort^8^. After quality control (QC) (see **Methods**), analyses for differentially methylated regions (DMRs) between the asthma/allergy cases and non-asthma/non-allergy controls were performed in the African American sample, the European American sample, and the combined sample (see **Methods** for additional details). Overall, we identified 16,611 DMRs that included 199,473 CpGs (**Table E4** in this article’s Online Repository).

### Selecting High-Value CpGs for an Asthma&Allergy Custom DNA Methylation Array

Our goal was to develop a custom array with 50,000 probes for high-value CpGs that would complement and serve as a “booster” for the EPIC array for studies of asthma and allergic diseases, as described above in **Methods** and in the Online Repository. After manufacture and array QC, 37,863 CpGs targeted by 42,192 probes remained (**Table E5** in this article’s Online Repository). The distributions of the 37,256 CpGs on the Custom array and the 789,290 CpGs on the EPIC arrays that passed final QC are shown by annotation category in **Fig. 2A**. Of note, 90% of CpGs on the Custom array overlapped with a transcription factor binding site (TFBS), and 94% overlapped with a predicted enhancer, compared to 56% and 54% of the EPIC array CpGs, respectively (Fisher exact test [FET] p<10^−16^ for both comparisons). Moreover, compared to EPIC CpGs, Custom CpGs were enriched in introns (52.8% vs. 47.2%) and exons (24.0% vs. 22.1%) and depleted in intergenic regions (23.1% vs. 30.7%) and 5’UTRs (6.8% vs. 7.4%) (FET p<10^−4^ for all comparisons) (**Fig. 2B**). The CpGs on the Custom array proportionally represented DMR-CpGs from the studies in URECA, COAST and combined analysis in WGBS and CpGs from the three categories of prior evidence (**Fig. E2** in this article’s Online Repository).

**Fig. 2.**
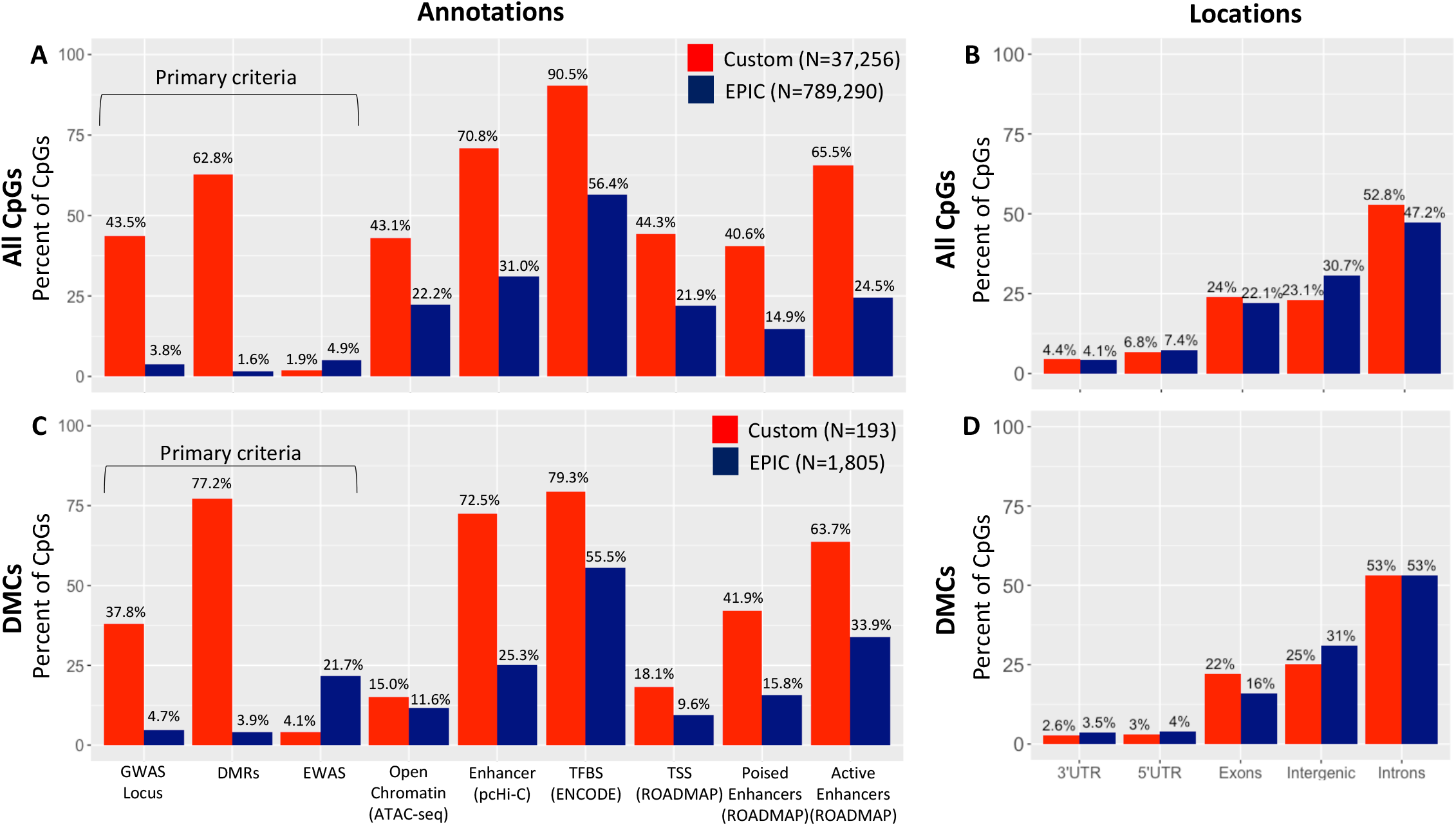
Proportions of Custom and EPIC CpGs by primary criteria, functional annotation category, and genomic location. All comparisons were performed with a Fisher’s Exact Test. **(A)** Proportions of Custom CpGs that passed processing QC in URECA on each array by three primary criteria and six functional annotation categories. CpGs on the Custom array were significantly enriched (p<2.2×10^−16^) in all categories compared to the EPIC array with the exception of prior EWAS, in which they were depleted on the Custom compared to the EPIC (p<2.2×10^−16^). **(B)** The distributions of CpGs by genomic location. Compared to the EPIC, CpGs on the Custom array were enriched in introns and exons and depleted in intergenic regions and 5’UTRs (p<10^−4^) but did not differ in any other categories. **(C)** Proportions of Custom and EPIC differentially methylated CpGs (DMCs) from each array by three primary criteria and six functional annotation categories. Compared to all CpGs, DMCs from both arrays were depleted at transcription start sites and in areas of open chromatin (Custom p=1.14×10^−7^ and p=4.11×10^−9^ ; EPIC p<2.2×10^−16^ or both, respectively). DMRs were marginally enriched for DMCs compared to all CpGs on the Custom array (p=0.066) but significantly enriched on the EPIC array (p=1.54×10^−6^). DMCs in prior EWAS studies of asthma and allergic diseases were modestly enriched on the Custom array (FET p=0.059) and significantly enriched on the EPIC array (p<2.2×10^−16^). In contrast, CpGs at GWAS loci for asthma and allergic diseases were not enriched among DMCs on the Custom array (p=0.32) and only modestly enriched on the EPIC array (p=0.042). (**D**) The distribution of DMCs by genomic location. The distribution of DMCs on both arrays did not differ from the distributions of all CpGs.

### Evaluating methylation patterns of CpGs on the Custom Array

To assess the reproducibility of methylation measures of CpGs on the Custom array, we compared methylation levels (measured as β values) in airway epithelial cell DNA in the WGBS to the same CpGs on the Custom and the EPIC arrays for the subset of individuals with all three (19 URECA subjects) or two (17 COAST subjects) measures. We included CpGs with at least 10 WGBS reads that overlapped with an array CpG that passed QC. This resulted in 502,229 CpGs for the WGBS-EPIC comparison in URECA and 24,744 and 20,861 CpGs for the WGBS-Custom array comparisons in URECA and COAST, respectively. The β distribution plots were similar and highly correlated between CpGs measured by both WGBS and an array (Spearman’s p<2.2×10^−16^ for each) (**Fig. E3** in this article’s Online Repository). Notably, however, compared to the EPIC array, CpGs on the Custom array were depleted for hypermethylated CpGs and enriched for intermediate methylated (IM) CpGs (β values 20-80%).

To more broadly assess EPIC and Custom array DNA methylation patterns, we compared the distributions of β values across different tissues using available data for the EPIC array in DNA from airway epithelial cells (this study), airway smooth muscle cells^24^, and buccal swabs, placenta cells, and cord blood^10^, and for the Custom array in DNA from nasal epithelial cells and nasal lavage cells from URECA children, and buccal cells, cord blood cells, or placenta tissue from infants in the VCSIP cohort^10^. The β distributions were similar across cell sources for the EPIC array (**Fig. 3A**) but showed varying patterns between cell types on the Custom array (**Fig. 3B**). Whereas an average of 68% (range: 60-79%) of CpGs on the EPIC array were either hypomethylated (0-20%) or hypermethylated (80-100%) in all six cell types, most CpGs on the Custom array (66% in nasal epithelial cells and 52% on average across cell types) were IM CpGs with depletions of β values between both 0-10% (24% in nasal epithelial cells and 29% on average) and 90-100% (10% in nasal epithelial cells and 20% on average). In fact, these β value distributions of CpGs on the EPIC array are remarkably similar to those in nine GTEx tissues (**Fig. E4** in this article’s Online Repository)^25^. To confirm that differences between the two arrays were a result of the selection criteria used for designing the array, we used the same criteria for the CpGs on the EPIC array. The 26,905 high-value EPIC CpGs also showed an enrichment for IM CpGs and a depletion of both hypomethylated and hypermethylated CpGs (**Fig. E5** in this article’s Online Repository).

**Fig. 3.**
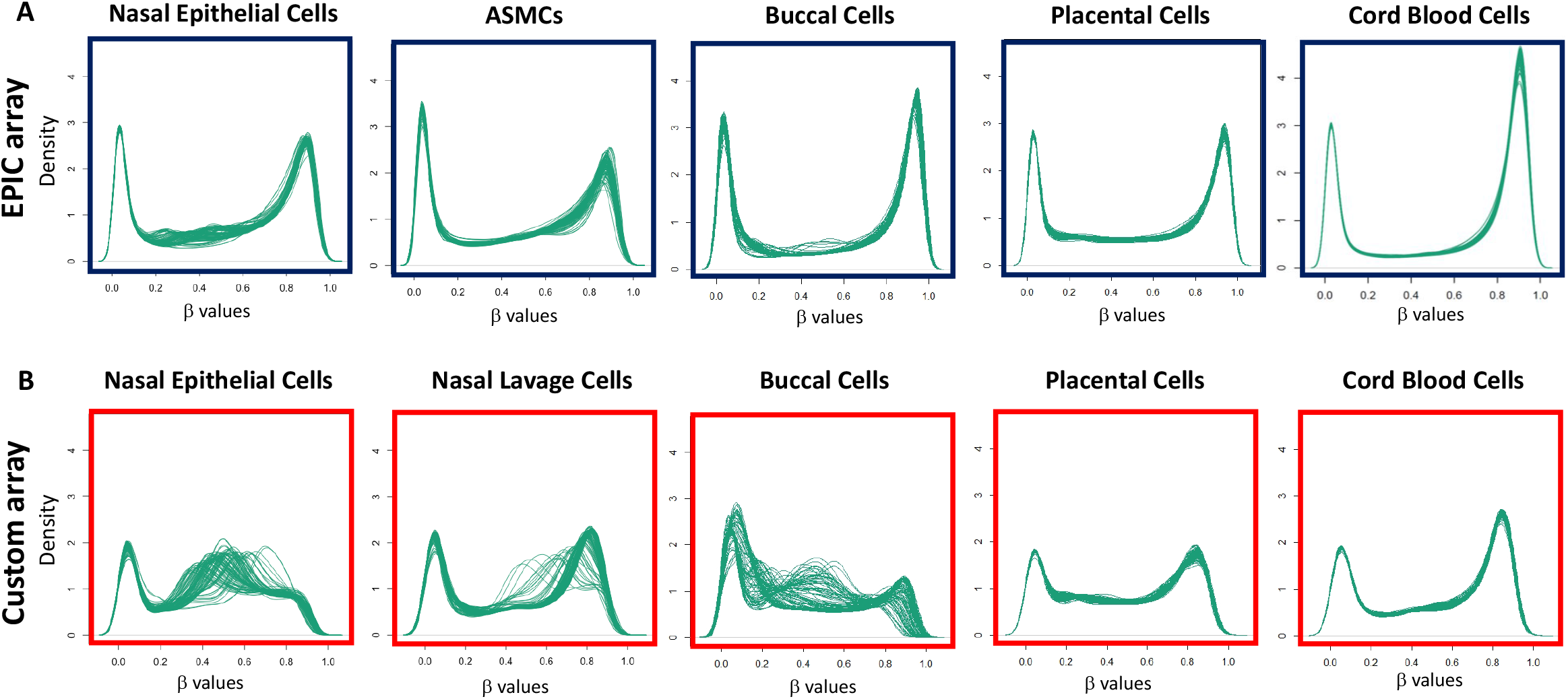
Cross-tissue comparisons of methylation levels for CpGs on the EPIC and Custom arrays. Density distribution plots of methylation levels, measured as β values, in 96 individuals after quantile (Custom) or quantile + SWAN (EPIC) normalizations (see **Methods**). The x-axis shows the proportion of methylation at CpG sites on the EPIC (**A**) 44 and Custom (**B**) arrays for individuals. All DNA methylation data were processed using the same pipeline. The nasal epithelial cell DNA (EPIC and Custom) and nasal lavage cell DNA (Custom) are shown for the same randomly selected URECA children. The buccal, placenta, and cord blood cells were from infants in the VCSIP Study.

### EWAS of Allergic Sensitization in 280 Multi-Ancestry Children using the Custom and EPIC Arrays

Our ultimate goal was to develop a DNA methylation array that could detect asthma- or allergy-associated differential methylation that is missed by the content on the EPIC array. AS is an IgE response to allergens^26^ and a crucial step in the development of allergic diseases and common phenotypes of asthma^21, 27, 28^. We conducted an EWAS of AS (see **Methods**) in airway epithelial cell DNA collected from 280 11-year old URECA children using the Custom and the EPIC arrays. The demographic and clinical description of the URECA children is shown in **Table I** and **Fig. E6** in this article’s Online Repository.

Following QC of the array, we performed two EWAS of AS in the same URECA children using the EPIC and Custom arrays (see **Methods**). Using a q-value threshold of 0.05, we identified 1,805 differentially methylated CpGs (DMCs) using the EPIC array and 193 DMCs using the Custom array (**Fig. 4A-B**), revealing an enrichment for AS DMCs on the Custom compared to the EPIC array (0.50% vs. 0.23% of CpGs on each array, respectively; FET p<2.2×10^−16^). Among the DMCs on the EPIC array, 295 (16%) were high-value EPIC CpGs, a significant enrichment compared to 3.5% of high-value CpGs among all CpGs on the EPIC array (FET p<2.2×10^−16^). In contrast to the different β distributions of CpGs on each array (**Fig. 3A**), the β distributions of DMCs were similar (**Fig. 4A-B**; middle panels), showing a near complete depletion of both hypomethylated and hypermethylated CpGs and an enrichment of IM CpGs among AS DMCs in airway epithelial cells. The DMCs on both arrays were distributed across the autosomes (**Fig. E7A-B** in this article’s Online Repository), however, Custom array DMCs showed spikes of association signals compared to the sparse distribution of EPIC array DMCs. In fact, 50% of DMCs on the Custom array were within 100bp of the next nearest DMC compared to only 3% of DMCs on the EPIC array (**Fig. 4D**). Regional clustering of DMCs on the Custom array provides greater confidence in and internal validation of EWAS results.

**Fig. 4.**
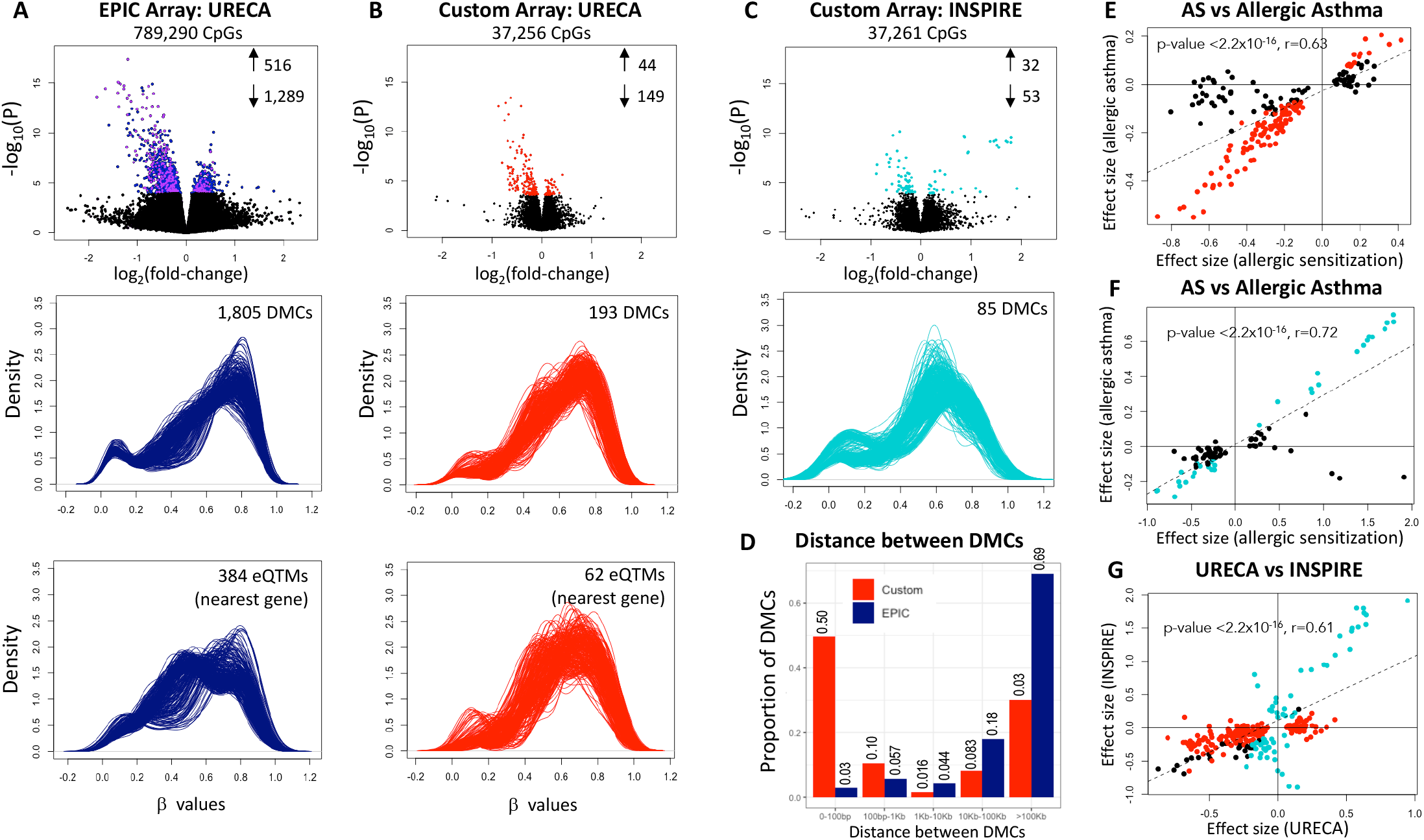
Allergic sensitization (AS) EWAS results in URECA and INSPIRE children using the EPIC and Custom arrays. **(A-C)** <u>Upper panel :</u> Volcano plots showing the log_2_-fold change in methylation (M-values) by proportion positive skin prick tests (x-axis) and the -log_10_(P-value) from the EWAS (y-axis). Significant DMCs at a q-value threshold of 0.05 are shown are shown in blue or purple (EPIC and high-value EPIC, respectively) in URECA, and Custom in red (URECA) or turquoise (INSPIRE). The number of AS DMCs that were hypermethylated or hypomethylated are shown as up and down arrows, respectively. (**A-C**) <u>Middle panel</u> : Density plots of the DMC β values in each EWAS. (**A-B**) <u>Lower panel:</u> Density plots of β values for DMCs that are eQTMs for their nearest genes (also see **Fig. E10** in this article’s Online Repository). **D**) Distributions of the distances of DMCs to the next nearest DMC on the Custom (red) and EPIC (blue) arrays. The distance to nearest DMC is shown on the x-axis; the proportion of DMCs in each distance bin is shown on the y-axis. (**E-F**) Correlation of effect sizes (log fold change) of AS DMCs and allergic asthma in URECA and INSPIRE. The effect size of DMCs for AS (x-axis) and allergic asthma (y-axis) are shown. Red or turquoise dots were associated with allergic asthma at a q-value ≤0.05 in URECA or INSPIRE, respectively; black dots are AS-only DMCs. (**G**) Correlation of effect sizes (log fold change) between DMCs identified in the INSPIRE AS 45 EWAS (x-axis) and the URECA AS EWAS (y-axis). Red or turquoise points are AS DMCs only in URECA or INSPIRE, respectively, at a q-value threshold of 0.05; black points are AS DMCs in both at a q-value ≤0.05. The dashed lines in E-G are the correlation lines.

Most of the functional annotation categories of DMCs were proportional to all CpGs on each array (**Fig. 2A** and **2C**; see **Fig. 2** legend for all statistical test results). However, DMCs from both arrays were depleted at transcription start sites and in areas of open chromatin compared to all CpGs on the arrays. Among the primary criteria (prior evidence), CpGs in DMRs were modestly enriched for DMCs compared to all CpGs on the Custom array and DMCs in prior EWAS studies were modestly enriched on the Custom array but significantly enriched on the EPIC array. In contrast, CpGs at GWAS loci were not enriched among DMCs on the Custom array and only modestly enriched on the EPIC array compared to all CpGs on the array. The genomic locations of the DMCs were proportional to all CpGs on the arrays (**Fig. 2B and 2D**).

Because AS is an important step in the development of childhood asthma, we next asked whether AS DMCs were also associated with allergic asthma. Considering only the AS DMCs on the EPIC array, 1,155 (64%) were also associated with allergic asthma at q-value <0.05 (1,805 tests). Among the AS DMCs on the Custom array, 115 (60%) were also associated with allergic asthma at q-value <0.05 (193 tests). The effect sizes were significantly correlated between AS DMCs and allergic asthma DMCs (r=0.68). Among the CpGs that were DMCs for both AS and allergic asthma, all showed the same direction of effect; among all AS DMCs, 93% showed the same direction of effect with allergic asthma (r=0.84) (**Fig. 4E**). The sample distributions of AS and allergic asthma are shown in **Fig. E8** in this article’s Online Repository.

### Validating the Custom Array in the INSPIRE Cohort

The URECA cohort is diverse with respect to ancestry (**Fig. E9** in this article’s Online Repository) but includes <1% non-Hispanic White children^7^. Therefore, to both replicate results of the EWAS in URECA children and assess the performance of the Custom array in a primarily non-Hispanic White population, we studied 5- to 7-year-old children from the INSPIRE cohort^9^. An EWAS was performed using the Custom array and nasal epithelial cell DNA from 474 children with measures of AS (see **Methods**). See **Table I** for demographic and clinical characteristics, **Fig. E6** in this article’s Online Repository for sensitization rates by allergen, and the Online Repository for details on processing and QC of the array data.

At a q-value threshold of <0.05 (37,261 tests), 85 CpGs on the Custom array were DMCs (0.2% of CpGs). The β distribution of the DMCs (**Fig. 4C**) and the patterns of associations (**Fig. E7C** in this article’s Online Repository) were similar to those observed in URECA. Among the AS DMCs in INSPIRE, 38% were also allergic asthma DMCs (q value<0.05 based on 85 tests), all with the same direction of effect. Among all AS DMCs in INSPIRE, 93% had the same direction of effect with allergic asthma (**Fig. 4F**). Moreover, among the AS DMCs in URECA, 25 (13%) were also AS DMCs in INSPIRE, which was a highly significant enrichment (p<2.2×10^−16^), and all AS DMCs had concordant directions of effect. Finally, the effect sizes of the 253 CpGs that were DMCs in either the URECA or INSPIRE AS EWAS were highly correlated (r=0.61; p<2.2×10^−16^) (**Fig. 4G**). The high reproducibility of EWAS results in two cohorts demonstrates that the high-value CpGs on the Custom array identified AS DMCs that are robust to ancestry, ascertainment strategies, age at sampling, and geography. The sample distributions of AS and allergic asthma are shown in **Fig. E8** in this article’s Online Repository.

### Evaluating the Biological Significance of DMCs in Airway Epithelial Cells

Because nearly all CpGs on the Custom array were selected to overlap with a TFBS or enhancer mark, we expected these CpGs to be correlated with the expression of genes in airway epithelial cells more often than CpGs on the EPIC array^29^. To test this, we used gene expression data^21^ in the same cells as those used for DNA methylation studies in 249 of the URECA children and defined two sets of genes from among the 15,551 genes detected as expressed in these cells: the nearest gene to each CpG and the promoter capture Hi-C (pcHi-C)-defined target gene in airway epithelial cells (see **Methods**). The latter identifies putative enhancers that physically interact with target gene promoters and account for the 3-dimensional structure of the genome that allows for distal regulatory elements to interact with and regulate the activity of promoters over large distances (e.g., up to 1 Mb or more). Among the nearest expressed genes to DMCs on the Custom (98 genes) and EPIC (1,449 genes) arrays, 63 were the nearest gene to CpGs on both arrays. Among the pcHi-C target genes for DMCs on the Custom (245 genes) or EPIC (1,155 genes) arrays, 95 were target genes of CpGs on both arrays. Thus, although no CpGs overlapped between the arrays, 101 of nearest or pcHi-C target genes were shared on both arrays (**Table E6** in this article’s Online Repository). The 318 nearest or pcHi-C target genes on the Custom array that were expressed in airway epithelial cells were enriched in 16 pathways (FDR <0.05), including “Th1 and Th2 cell differentiation,” “JAK-STAT signaling,” “Th17 cell differentiation,” and “Viral protein interaction with cytokine and cytokine receptor” (**Table E7** in this article’s Online Repository and Supplementary **Methods** in the Online Repository). In contrast, the 2,366 nearest or pcHi-C target genes on the EPIC array that were expressed in airway epithelial cells were not enriched for any pathways (smallest FDR = 0.13). These findings further demonstrate that the CpGs on the Custom array are enriched in pathways relevant to asthma and allergic disease.

For each CpG-gene pair, we tested for correlation between DNA methylation and expression levels of their nearest and pcHi-C target gene(s) to identify eQTMs. Significantly more CpGs on the Custom array were eQTMs with their nearest gene (23%) or pcHi-C target gene (16%) compared to CpGs on the EPIC array (11% and 9% respectively; FET p<2.2×10^−16^ in both analyses) (see the Online Repository). The high-value EPIC CpGs were enriched for eQTMs compared to all EPIC CpGs (20% and 12%, respectively; FEΤ p<2.2×10^−16^ in both analyses). Although more AS DMCs were eQTMs compared to non-DMCs on both arrays, significantly more were on the Custom compared to the EPIC array (nearest gene 35% vs. 20% [FET p = 0.0019] and pcHi-C 22% vs 15% [FET p = 0.0082], respectively) (**Table E8** in this article’s Online Repository). The β distributions for DMCs on the Custom and EPIC arrays that were eQTMs were further enriched for IM CpGs and depleted of CpGs at the extremes of the distribution (**Fig. 4** and **Fig. E10** in this article’s Online Repository).

### Further Insights into the Epigenetic Regulation of Gene Expression at EWAS Loci

To examine the DMCs and their associations with gene expression more closely, we generated regional association plots for the 10 most significant DMCs in the URECA EWAS using the EPIC and Custom arrays. The 10 most significant EPIC DMCs were at 10 loci (**Fig. E11** in this article’s Online Repository). Seven were “solitary” with no other DMCs within 500kb and three had one other DMC within 6.5kb. Among the solitary DMCs, six were high-value EPIC CpGs and all 10 were reported as DMCs in airway epithelial cells in previous EWAS of asthma or allergic phenotypes. Six of the 10 DMCs were within genes and four were intergenic. CpGs from the Custom array were present in four of these regions, but none were AS DMCs. In contrast, the 10 most significant Custom DMCs were at six loci and only two were solitary (**Fig. E12** in this article’s Online Repository). One solitary DMC in *ALOX15* was near a high-value and a non-high-value EPIC DMC; the other solitary CpG in the PDE6A gene was also a DMC in the INSPIRE EWAS. The other four loci had spikes of association, often with a combination of DMCs from both arrays in URECA and the Custom array in both URECA and INSPIRE. Eight of the 10 most significant DMCs were within genes; two were intergenic.

We selected three additional regions as examples of patterns of associations (**Table E9** in this article’s Online Repository). The first includes a spike of 11 Custom array DMCs in URECA, 12 in INSPIRE and one high-value EPIC DMC in exons 2 and 3 of the *CISH* (Cytokine Inducible SH2-Containing Protein) gene, a member of the SOCs family of negative regulators of cytokine signaling (**Fig. 5A**). There was one nearby DMC from the EPIC array downstream of *CISH*. The lead Custom and EPIC URECA DMCs were eQTMs for *CISH*, with increased methylation associated with decreased gene expression and fewer allergic sensitizations. The EPIC high-value CpG at this locus was identified in previous asthma/allergy EWAS^30, 31^, but this locus has not been associated with asthma or allergic diseases in GWAS. *CISH* expression is induced by IL-13 in bronchial epithelial cells and macrophages^32^, and has been implicated in eosinophil physiology^33^ and eosinophilic inflammation^32^. Our studies are consistent with these findings and further indicate that increased expression of *CISH* is associated with increased sensitization to allergens and that the regulation of *CISH1* expression may be epigenetically mediated.

**Fig. 5.**
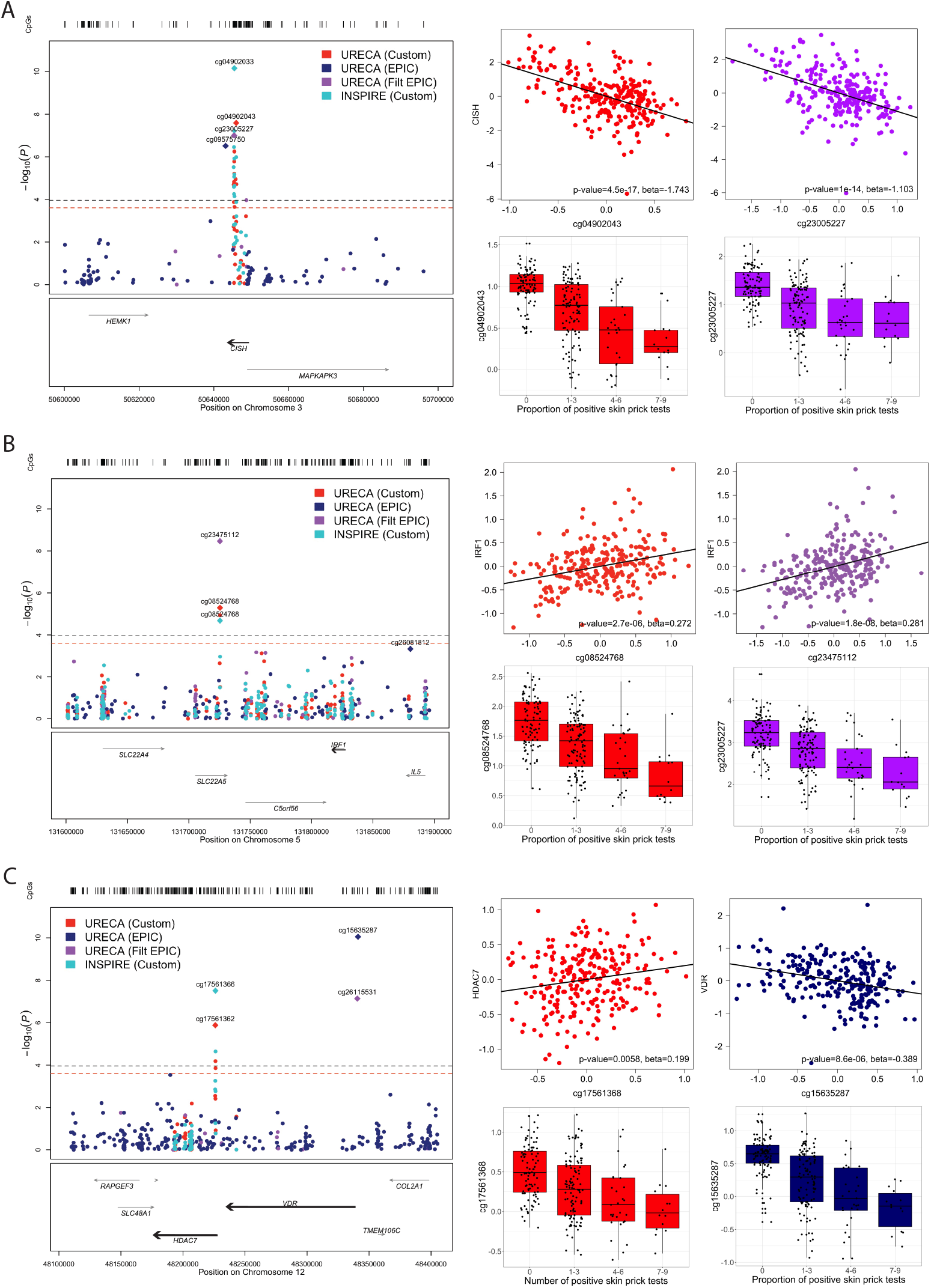
DMCs and eQTMs at 3 exemplary loci. In each panel, locus zoom plots show the DMCs from the three AS EWAS using the EPIC array (URECA; blue points = non-high value CpGs; purple point = high-value [filtered] CpGs) and Custom array (URECA, red points; INSPIRE, turquoise points). The genomic locations and genes are shown on the x-axis; EWAS p-values are shown on the y-axis. The dashed horizontal lines show the 0.05 q-value threshold for the Custom array in URECA (red) and for the EPIC array in URECA and Custom array in INSPIRE (blue and purple overlaid). The bar code at the top of the plot shows the location of CpGs at this locus on both arrays. The most significant DMC in the region for a given EWAS (cohort and platform; see legend) is illustrated by a diamond; additional DMCs appear as circles. Correlations between methylation levels (x-axis) of the lead Custom and high-value EPIC CpGs in URECA and expression of relevant genes (y-axis) in epithelial cells from URECA children are shown in the upper right of each panel, and boxplots of the proportion of positive skin prick tests (x-axis) and methylation levels for the lead Custom and high-value EPIC CpGs in URECA (from the EWAS) are shown in the lower right of each panel. **A)** *CISH* locus on chromosome 3. **B)** *SLC22A5*-*IRF1* locus on chromosome 5. **C)** *HDAC7-VDR* locus on chromosome 12.

A second locus included one Custom DMC in URECA and INSPIRE, a second Custom DMC in URECA, and one high-value EPIC DMC in an intron of *SLC22A5* (Solute Carrier Family 22 Member 5) (**Fig. 5B**). The lead DMC was a high-value EPIC CpG, which was also identified in prior EWAS in nasal epithelium^30, 31^. None of the DMCs were eQTMs for *SLC22A5*, but all three were in a region that physically interacted with the promoter of Interferon Response Factor 1 (*IRF1*), 91kb away, and were eQTMs for *IRF1* (p=2.4×10^−7^, beta=0.27 and p=2.7×10^−6^, beta=0.27). This region is at a GWAS locus for adult- and childhood-onset asthma and hay fever^34, 35^ (**Table E9** in this article’s Online Repository), possibly with sex-specific effects^36^. Genetic variation in the *IRF1* gene has been associated with increased expression of pro-inflammatory genes and IL-13 secretion in peripheral blood cells^37^. Our studies further demonstrate long-range epigenetic regulation of *IRF1* expression in airway epithelial cells, with increased methylation levels associated with increased gene expression and decreased numbers of sensitizations.

At a third locus, we observed a spike of three Custom array DMCs in URECA and two in INSPIRE upstream of the HDAC7 (Histone Deacetylase 7) gene (**Fig. 5C**). One Custom DMC (upstream of *HDAC7* and downstream of *VDR*) was an eQTM for *HDAC7*. Histone deacetylases (HDACs) have diverse functions, including regulation of inflammatory genes^38^, and impaired barrier function in asthmatics was both induced by HDAC activity and reversed by inhibition of endogenous HDAC^39^. In this study, increased methylation in the *HDAC7* gene was associated with increased expression of *HDAC7* and fewer sensitizations. At this extended locus, two EPIC DMCs were in an intergenic region 42kb upstream of the *VDR* (Vitamin D Receptor) gene. Vitamin D deficiency in childhood has been associated with increased risk for persistent asthma^40^ and high dose vitamin D supplementation in pregnancy reduced risk of recurrent wheeze in childhood^41^. Here, increased methylation (EPIC array) near the *VDR* gene was associated with decreased VDR expression and sensitization to fewer allergens. The *HDAC7-VDR* region has been identified in GWAS of childhood- and adult-onset asthma^34^. This locus demonstrates the complementarity of the Custom and EPIC arrays in detecting differential methylation relevant to asthma and allergic diseases.

## Discussion

The epigenome plays a critical role in regulating gene expression in a context-specific manner, such as in specific cell types or in the presence of environmental exposures or disease states. DNA methylation, for example, is responsive to disease promoting exposures in both *in vitro* cell models^24, 42-44^ and *ex vivo* cells from individuals who are healthy and with disease^1, 45-48^. Yet the DNA methylome has been underexplored and largely limited to the CpGs on commercial arrays. As a result, it is unknown whether the 800,000 or fewer CpGs interrogated in nearly all EWAS to date include CpGs most relevant to any specific exposure, disease, or cell type. Here, we addressed this question by designing a custom Allergy&Asthma DNAmethylation array with 37,863 high-value CpGs that are not on the commercial EPIC array. Our study design allowed us to make important observations regarding features of high-value CpGs and to propose a pipeline that can be used to prioritize CpGs from among the more than 28 million CpGs in the human genome.

Although we did not use methylation levels as a selection criterion, the CpGs selected were significantly enriched for IM CpGs (β values 20-80%). The CpGs that were DMCs and eQTMs were further enriched for IM CpGs on both the EPIC and Custom arrays, suggesting that this feature is a signature of high-value, variable and likely functional CpGs in the genome. In contrast, the non-IM CpGs may be more functionally constrained and less responsive to environmental perturbations. Our results are supported by previous WGBS studies showing that IM CpGs were more likely to be tissue specific, to vary between individuals, and to play important roles in gene regulation^29, 49^. Our study further revealed that CpGs with intermediate levels of methylation are also more likely to be associated with AS, an important clinical phenotype that reflects both an immune response to past allergen exposures^26^ and a risk factor for the development of asthma and allergic diseases^21, 27, 28^. Although the CpGs on the custom array were overall enriched for IM CpGs, AS DMCs were further enriched, with a depletion of very hypomethylated (0-20% methylation) and very hypermethylated (80-100%) CpGs. This important observation was further supported by EWAS results using the EPIC array. Whereas the EPIC array is enriched for hypomethylated and hypermethylated CpGs in all tissues, the AS DMCs from the EPIC were depleted for CpGs at both extremes and enriched for IM CpGs, similar to the DMCs on the custom Allergy&Asthma array. We further showed that the 26,905 high-value EPIC CpGs were also enriched for IM CpGs and enriched among AS DMCs: 16.3% of the DMCs in the AS EWAS were among the high-value EPIC CpGs compared to 3.4% of all EPIC CpGs. These results, combined with previous studies^29, 49^, could justify filtering CpGs on the EPIC array to include just IM CpGs (β=20-80%) to enrich for functional CpGs and increase power to detect DMCs associated with exposures or disease outcomes. As proof-of-concept, including only IM EPIC CpGs and using a q-value of 0.05, 2,182 CpGs were AS DMCs (0.70% of all IM CpGs), more than three times the proportion of DMCs among all CpGs (1,805 AS DMCs; 0.23% of all EPIC CpGs). Finally, DMCs that were eQTMs were further enriched for IM CpGs (**Fig. 4** and **Fig. E10** in this article’s Online Repository), supporting their role in gene regulation.

In addition to the enrichment of IM CpGs on the Custom compared to the EPIC array, the different patterns of associations with AS were notable. Whereas DMCs on the EPIC were more sparsely distributed throughout the genome, the DMCs on the Custom often showed spikes of associations, analogous to GWAS peaks. The spatial organization among DMCs on the Custom array provides confirmatory evidence for their veracity, which is generally not available for EPIC CpGs (Figure 4D). Another distinguishing feature of the DMCs on the Custom array was their enrichment in exons and depletions in intergenic regions compared to DMCs on the EPIC array. This is consistent with results of an earlier study that observed enrichments of IM CpGs in exons, where exon expression at those sites correlated with DNA methylation^29^.

The CpGs included on the custom Allergy&Asthma array were selected based on prior evidence of associations with asthma or allergic diseases using three classes of evidence: within a DMR in our WGBS study, in a previous published EWAS, or at published GWAS loci. Overall, 62% of CpGs on the custom array were selected from DMRs compared to 1.6% of all CpGs on the EPIC array, yet the DMR CpGs were enriched among DMCs on both the custom (77.2%) and EPIC (3.9%) arrays. In contrast, few previous EWAS CpGs were on the Custom array because we excluded all CpGs on the EPIC from the Custom array, yet we still observed a modest enrichment of EWAS CpGs among Custom array DMCs (1.9% overall vs. 4.1% of DMCs) and a highly significant enrichment among DMCs on the EPIC array (4.9% overall vs. 21.7% of DMCs). These represent replications of results from previous EWAS of asthma and allergic phenotypes in the URECA children. Surprisingly, CpGs at asthma and allergy GWAS loci were only modestly enriched among EPIC DMCs and not at all enriched among the Custom DMCs, even though the EWAS in URECA and INSPIRE revealed many examples of AS DMCs at important GWAS loci. The lack of enrichment of Custom array CpGs at GWAS loci among DMCs may reflect the less direct evidence for DNA methylation mechanisms at GWAS loci compared to CpGs from previous EWAS or DMR studies, the large proportion of CpGs at GWAS loci included on the Custom array (44% of all CpGs), and the enrichment of AS DMCs among genic regions whereas GWAS loci are more often in intergenic regions.

Our study has limitations. First, due to cost and technical limitations, the Custom array included only 37,863 CpGs out of the 92,024 high-value CpGs that we identified with our pipeline. Therefore, together with the 26,905 high-value EPIC CpGs, we only surveyed 54% of potential high-value CpGs in airway epithelium that are relevant to asthma and allergic diseases. The next generation (v1.1) of the Custom array includes ∼8,000 more high-value CpGs, reducing this gap, although alternative array designs or other **methods** that allow the inclusion of the 118,929 high-value CpGs would be a powerful tool for assessing epigenetic mechanisms in these conditions. Second, we have not yet evaluated the performance of this array in blood cells, a much more easily sampled tissue. Those studies are currently underway. Lastly, we did not functionally validate the correlations between CpGs and gene expression or identify the “causal” CpG(s) driving the correlations, which should be considered in follow-up studies of any of the EWAS regions reported here.

In summary, our study revealed high-value CpGs in airway epithelial cells that are not interrogated on the EPIC array. The allergic sensitization EWAS revealed potential epigenetic regulatory mechanisms both for genes previously implicated in asthma or allergy pathogenesis but not identified in GWAS (e.g., *CISH* ^32, 33^, *NEDD4L* ^50, 51^, *PRDM10* ^52, 53^) as well as for genes at replicated GWAS loci (e.g., *IRF1, CLEC16A, HDAC7/VDR, ALOX15, SLC22A5*) (**Table E9** in this article’s Online Repository). These associations were robust to race/ethnicity, ascertainment, age, and geography. We suggest that similar pipelines can be used to identify high-value CpGs for other disease groups or specific environmental exposures leading to the discovery of currently invisible portions of the epigenome that are relevant to human health.

## Supporting information

Online Repository

## Data Availability

All data produced are currently being made available at GEO and dbGaP.

## Abbreviations

AS: allergic sensitization
DMC: differentially methylated CpG
DMR: differentially methylated region
eQTM: expression quantitative trait methylation
EWAS: epigenome-wide association study
FET: fisher exact test
GWAS: genome-wide association study
IM: intermediate methylation
LF: latent factors
NEC: nasal epithelial cells
PCA: principal components analysis
SPT: skin prick test
WGBS: whole-genome bisulfite sequencing

## Acknowledgments

The authors wish to thank our ECHO colleagues, the medical, nursing, and program staff, and the children and families participating in the ECHO cohorts. We also acknowledge the contribution of the following ECHO program collaborators: ECHO Components – Coordinating Center: Duke Clinical Research Institute, Durham, NC: Smith PB, Newby KL.

## Author Contributions

A.M., E.E.T., B.A.H. performed all data analyses; A.M., E.E.T., B.A.H., C.G.M., and C.O. designed the study and wrote the manuscript; P.F. oversaw the laboratory assays; L.E.S-K., T.G., L.B.B., M.K., G.T.O., K.R.S., R.A.W., C.T.M., E.R.S., T.H., D.J.J., and J.E.G. provided clinical samples and information and consulted on study design; M.C.A. provided gene expression data; K.C.B., R.A.M., and K.H. consulted on study design. All authors read, commented on and approved the manuscript.

## Data Sharing Plans

The gene expression data used this study are available on the Gene Expression Omnibus (GEO) repository (accession number GSE145505). The DNA methylation array data used in the EWAS study are available in GEO (accession number in progress). The whole-genome bisulfite sequencing data are available in dbGaP (in progress).

