## Supplementary material for "A Functional Genomics Pipeline to Identify High-Value Asthma and Allergy CpGs in the Human Methylome": Online Repository

**This PDF file includes:**

- Supplementary Methods
- Figures E1 to E12
- Tables E1 to E9
- Supplementary References

**Other online repository materials for this manuscript include the following:**

- Supplementary Dataset 1 and Supplementary Dataset 2

| Table of contents | | Page |
|---|---|---|
| Supplementary Methods | | 2 |
| **Supplementary Figures** | | |
| Fig. E1 | Overview of study design and CpG selection criteria | 12 |
| Fig. E2 | Overlap of CpGs by prior evidence and WGBS source | 14 |
| Fig. E3 | Comparison of β values between WGBS data and Custom and EPIC arrays | 15 |
| Fig. E4 | Beta distribution plots from nine GTEX tissues | 16 |
| Fig. E5 | Beta value distribution of the high-value EPIC compared to the full EPIC array | 17 |
| Fig. E6 | Allergic sensitization in URECA and INSPIRE | 18 |
| Fig. E7 | Manhattan plots illustrating EWAS results of allergic sensitization | 19 |
| Fig. E8 | Overlap of allergic sensitization and allergic asthma cases and controls | 20 |
| Fig. E9 | Ancestry PCA plot of self-identified race/ethnicity of 280 URECA subjects | 21 |
| Fig. E10 | Density plots showing β value distributions among eQTMs and their nearest genes and pcHi-C target genes for the EPIC (A) and Custom (B) arrays | 22 |
| Fig. E11 | Regional association plots for the 10 most significant DMCs in the URECA EWAS using the EPIC array | 23 |
| Fig. E12 | Regional association plots for the 10 most significant DMCs in the URECA EWAS using the Custom array | 24 |
| **Supplementary Tables** | | |
| Table E1 | Published EWAS studies of asthma and allergic disease | 25 |
| Table E2 | GWAS studies of asthma and allergic disease used to define regions for CpG inclusion | 26 |
| Table E3 | PCA QC of DNA methylation data and identifying batch effects | 27 |
| Table E4 | Description of DMRs identified in the WGBS study | 29 |
| Table E5 | CpGs included on the Custom array by annotation category | 32 |
| Table E6 | Nearest and pcHi-C target genes to DMCs across arrays | 33 |
| Table E7 | Results of KEGG pathway analysis on genes nearest DMCs and pcHi-C target genes on the Custom array using iPathway Guide | 41 |
| Table E8 | Enrichment of eQTMs among all CpGs on the Custom array and among DMCs | 42 |
| Table E9 | Annotation information for the three selected loci | 43 |
| **Supplementary References** | | 44 |

**Supplementary Methods**

Cohorts Included in the WGBS or EWAS Studies

URECA: The URECA study is an observational birth cohort study initiated in 2005 in Baltimore, Boston, New York City, and St. Louis under the NIAID-funded Inner City Asthma Consortium [E1]. Either the pregnant mother or the father of their unborn child had a history of asthma, allergic rhinitis, or eczema. Asthma was assessed at age 10 according to a definition that considered symptoms, diagnosis by a health care provider, and measurements of pulmonary function, as described [E2]. Skin prick testing was performed at age 10 (255 subjects) or age 7 (25 subjects) and included the following allergens: mouse epithelia, dog epithelia, *Dermatophagoides fainae* (mite), *Dermatophagoides pteronyssinus* (mite), cat hair, rat epithelia, American/German cockroach mix, German cockroach, *Alternaria tenuis* (mold), *Aspergillus* mix, ragweed mix, tree pollen (oak or birch), *Penicillium Notatum/Penicillium Chrysogenum*, and Timothy grass. Twenty African American children were selected for the WGBS studies (10 with asthma and allergic disease [3 females, 7 males], 10 without asthma or allergic disease [3 females, 7 males]), and 280 unrelated children were included in the EWAS using both the Custom and EPIC arrays.

COAST: The COAST study is an observational birth cohort study initiated in 1998 in Madison, Wisconsin [E3]. Pregnant women were recruited in the third trimester of pregnancy if the mother or the father had a history of asthma or allergic diseases. Asthma was assessed beginning at age 6 years. Children were diagnosed with asthma if they fulfilled at least one of the following criteria: (1) physician-diagnosed asthma; (2) frequent albuterol use for coughing or wheezing episodes as prescribed by a physician;

(3) use of a prescribed daily controller medication; (4) an implemented step-up plan, including use of albuterol or inhaled corticosteroids during illness as prescribed by a physician; (5) use of prednisone for an asthma exacerbation. Twenty participants of European American ancestry were selected for the WGBS studies (10 with asthma and allergic disease [5 males, 5 females], 10 without asthma or allergic disease [5 males, 5 females]).

INSPIRE: The INSPIRE study is a population-based observational birth cohort of healthy infants in central Tennessee [E4]. Infants were enrolled following delivery into this population-based longitudinal study in central Tennessee. Children were followed longitudinally for the outcomes of asthma and allergic diseases. Subjects were tested for the following thirteen allergens: dog, cat, *Dermatophagoides pteronyssinus* and *Dermatophagoides farinae* mix, American/German cockroach, *Penicillium Notatum*/*Penicillium Chrysogenum*, *Alternaria Tenuis*, *Cladosporum Herbarum*, *Aspergillus* mix, Ragweed mix, Eastern 6 Tree mix, K-O-T Grass mix, Maple/Box Elder mix, and Weed mix.

DNA Extraction protocols:

URECA: Nasal brushings were obtained at age 11 years, as described [E2]. Total DNA was isolated from brushes stored in RLT Plus lysis buffer. Samples were thawed, vortexed, and then spun to collect the supernatant, which was transferred to fresh tubes. Seventy percent EtOH was used to wash the brushes and original tubes, which was then transferred to the new tubes. The samples were spun through a Qiashredder column (Qiagen) and then extracted using AllPrep DNA/RNA mini kits (Qiagen) with 100ul elution volumes for DNA following the manufacturer's protocol. Nasal lavage DNA was isolated using the QIAamp DNA Micro Kit. COAST: DNA was extracted from nasal brushings obtained at ages 18-20 years, as described [E5]. INSPIRE: Flocked nasal swabs were obtained at age 5-6 years and samples were stored at –80°C in RNA lysis buffer. RNA and DNA were isolated from the swabs using the Qiagen AllPrep DNA/RNA kit. VCSIP: Placental DNA was extracted from powdered tissue under liquid nitrogen, using the QIAcube for automated nucleic acid extraction. Cord blood DNA was extracted from 200µL of whole blood using the QIAamp DNA Mini Kit and the QIAcube, and buccal DNA was extracted from buccal swabs following proteinase K digestion using the Maxwell 16 Blood DNA Extraction Kit and the Maxwell 16 Instrument for automated nucleic acid extraction.

DMR Analysis

DMR analyses were conducted in the African American sample (n=20), the European American sample (n=19), and the combined African American and European American sample (n=39). The methylation data were then smoothed using BSmooth [E6], and DMRs were called using the bsseq package (version 1.14) [E6]. Only CpGs covered by at least 10 reads in 80% of the cases and controls (AA only, EA only, and combined n=39) were included. T-statistics cutoffs were based on 5% quantiles, and a maximum gap of 300bp was required between CpGs to define a cluster, as recommended by BSmooth. To maximize the number of DMRs, we required three or more CpGs per DMR and a minimum of 5% difference in methylation levels between the allergy asthma cases and non-allergy, non-asthmatic controls. The union of DMRs between analyses was assessed using the *reduce* function from the GenomicRanges R package (version 1.30).

Selection of CpGs for Custom Array

Our pipeline consisted of three steps. In the first step, we prioritized regions with prior evidence of association with asthma or allergic diseases (atopic dermatitis/eczema, allergic rhinitis/hay fever, and food allergy) from three categories of studies. The first category included the 199,473 CpGs within the DMRs from the WGBS. The second category included CpGs from previous DNA methylation studies (EWAS) of asthma or allergic diseases. For this, we conducted a literature search for array-based studies of DNA methylation in asthma and allergic disease. We identified 15 studies that conducted 25 EWAS of asthma- or allergy-related phenotypes, five of which were conducted in respiratory epithelium and 10 in blood (**Table E2**). We also included CpG sites for five additional genes from two candidate-gene DNA methylation studies of food allergies; *FOXP3* [E7] and *IL4*, *IL5, IL10* and *INFG* [E8]. In total, 19,057 unique DMCs were identified. The third category included CpGs located within the 140 GWAS loci defined in two recent large studies of adult-onset and childhood-onset asthma [E9] and allergic diseases (asthma, hay fever and eczema) [E10] in UK Biobank subjects [E11], which included nearly all the loci reported in other GWAS in multi-ancestry populations. In total, the GWAS loci covered a total of 570,350 CpG motifs. We also included CpGs for the *MALT1* gene that was significantly associated with peanut allergy in a genome-wide interaction study [E12]. We next removed duplicate CpGs from among the three categories of prior studies, CpGs on the EPIC array, CpGs in ENCODE blacklist regions [E13], and those in which the cytosine nucleotide overlapped with common SNPs (MAF > 5%) in 1000 Genomes CEU or YRI populations (**Table E5**). A total of 696,225 CpGs remained for consideration in the second step.

To further prioritize the CpGs, in the second step we considered their overlap with six functional annotations: 1) ENCODE [E14] TFBSs from all cell types; 2-4) ROADMAP Epigenetics [E15] transcriptional start sites, poised enhancers and active enhancers from smooth muscle (E078, E076, E103, E111), epithelial (E055, E056, E059, E061, E058), and blood cells (E062, E034, E045, E044, E043, E039, E041, E042, E040, E037, E048, E038, E047, E029, E050, E032, E046); 5) ATAC-seq in human cultured bronchial epithelial cells exposed to rhinovirus or a vehicle control from asthmatic and non-asthmatic individuals [E16], and 6) pcHi-C from *ex vivo* human bronchial epithelial cells [E16]. We required that CpGs at DMRs overlapped with at least three functional annotations or prior evidence (GWAS or EWAS), CpGs from previous EWASs overlapped with at least one functional annotation or prior evidence (GWAS or DMR), and CpGs at GWAS loci overlapped with at least four functional annotations or prior evidence (EWAS or DMR). Lastly, we selected all remaining CpGs that were within both a GWAS locus and a DMR.

Of the 92,024 resulting high-value CpGs identified, 53,700 were amenable to the Illumina design algorithm, and 44,047 CpGs targeted by 49,999 probes were selected by us for manufacturing. Fourteen percent of the CpGs failed manufacturing, resulting in a Custom array with 38,541 CpGs targeted by 43,605 probes. After removing probes with unknown origins and those without genomic coordinates, 37,863 CpGs passed array QC. The locations of this final set of CpGs, their selection among the three primary criteria, and their overlap with the annotation categories are shown in Supplementary Dataset 1.

Finally, we removed probes that included a common SNP (MAF≥5%) within 3 bp of the CpG interrogation site, probes that failed the minfi QC, and probes on the X chromosome, leaving 37,256 CpGs following both array and processing QC.

Processing Custom and Epic array DNA methylation data in URECA

For the EPIC array, probes that failed (detection $P < 0.01$ in at least 25% of samples), overlapped with known SNPs with MAF of at least 5% in African American or European Americans, mapped to the X or Y chromosomes, overlapped ENCODE blacklist regions, or mapped to multiple locations in a bisulfite-converted genome were removed. Raw probe values were background corrected using preprocessIllumina(bg.correct="TRUE", normalize="no"), and quantile normalization was performed using ENmix (v 1.30.01) (https://doi.org/10.1186/s13148-021-01207-1), followed by SWAN [E17] normalization. Samples that failed sex checks using the getSex function in minfi were removed. DNA concentration, collection site, array and plate showed batch effects by principal components analysis (PCA) [E18]. The effects of collection site, array, and plate were removed using ComBat [E19]; DNA concentration was removed using linear regression. We estimated unobserved variation in the data correlated with our phenotype of interest [E20] and included these latent factors in the model [E20]. The first three ancestry PCs were also included as covariates to capture the effects of admixture in the sample. After QC, we retained 789,290 (91.1%) of CpGs on the EPIC array for analysis.

For the Custom array, probes that failed (detection $P < 0.01$ in at least 25% of samples), contained a SNP with MAF of at least 5% in African American or European

Americans within 3bp of the CpG interrogation site, mapped to the X chromosome, or were missing genomic coordinates were removed. Raw probe values were background corrected using preprocessIllumina(bg.correct="TRUE", normalize="no"), and quantile normalization was performed using ENmix. Additional probe-type normalization using SWAN was not applied because the dye intensities for the methylated and unmethylated type I and type II probes had very different distributions. DNA concentration, collection site, array, and plate showed batch effects by PCA. The effects of site, plate, and chip were removed using ComBat, and DNA concentration was removed using linear regression. We estimated unobserved variation in the data that was not correlated with the phenotype of interest and included these latent factors in the model. To account for regional differences in the density of Custom array CpGs on the estimation of latent factors, we used the partition.params function in FALCO [E21] (https://github.com/chrismckennan) to partition groups of CpGs into independent units (default size = 100 kb). The first three ancestry PCs were included as covariates to capture the effects of admixture in the sample for analyses in URECA. After QC, we retained 37,256 (98.1%) CpGs on the Custom array for analyses.

For comparison purposes, we filtered the CpGs on the EPIC array through the same pipeline described above for selecting CpGs for the Custom array; we refer to these as “high-value” EPIC CpGs. Of the 789,290 CpGs on the EPIC array, 26,905 (3.4%) were considered high-value, similar in number to the 37,863 CpGs that passed QC on the Custom array (Supplementary Dataset 2).

Processing Custom and Epic array DNA methylation data in INSPIRE

The same pipeline used in URECA was used to process INSPIRE Custom methylation data. Twenty-one probes failed *P*-value detection and were removed from the analysis. We estimated sex using the X chromosome CpGs and removed five samples with discrepancies between these classifications and reported sex. DNA concentration, collection year, plate, and array were identified as having batch effects by PCA (**Table E3**). The effects of collection year, plate, and array were removed using ComBat, and DNA concentration was removed using linear regression. Because DNA concentration was correlated with other variables, it was also included as a covariate in the model, along with sex, parent-reported race/ethnicity, and latent factors estimated using the same methods described above for URECA. A total of 37,261 CpGs passed processing QC and were included in the analysis.

Correlations of methylation level with expression of nearest gene and target promoter capture Hi-C gene

Correlations between methylation levels at each CpG with the expression levels of the nearest gene and the pcHi-C target genes [E16] were assessed using linear regression in both the full data set (all CpGs that passed QC on each array) and in the DMCs. Comparisons were only assessed for DMCs falling in "capture end regions" (± 1kb) that interacted with gene promoters [E16]. Sex, ancestry PCs, and epithelial cell composition were included as covariates in the model. The nearest gene was annotated using the GenomicRanges package v1.46.1 in R with the gene list from the R biomaRt package v2.50.2 using ensemble GRCh37. A false discovery rate of 5% was used to assess significance.

Pathway analysis

Pathway over-representation testing was performed with Advaita Bio's iPathwayGuide (https://www.advaitabio.com/ipathwayguide) using KEGG Pathways (release 100.0+/11-12, Nov 2021). For the Custom and the EPIC arrays, the 318 and 2,366 genes, respectively, that were either the nearest or pcHi-C target gene to a DMC, were entered as significant against a background of all nearest genes to CpGs on each arrays that are expressed in NECs (n=14,049).

Estimating genetic ancestry

Ancestry PCs were estimated in the URECA children using a set of 3,534 SNPs that were genotyped in URECA and in reference panels from the 1000 Genomes Project (1KG; n=156)[E22] and the Human Genome Diversity Project (HGDP; n=52). European, West African, and East Asian reference samples were randomly selected from CEU (n=52), YRI (n=52), JPT (n=26), and CHB (n=26) samples, respectively, in the phase 3 1KG reference panel [E22]. Ancestry PCs were calculated using PC-Air [E23]. The first two ancestry PCs for the URECA children in our study are shown in **Fig. E9.**

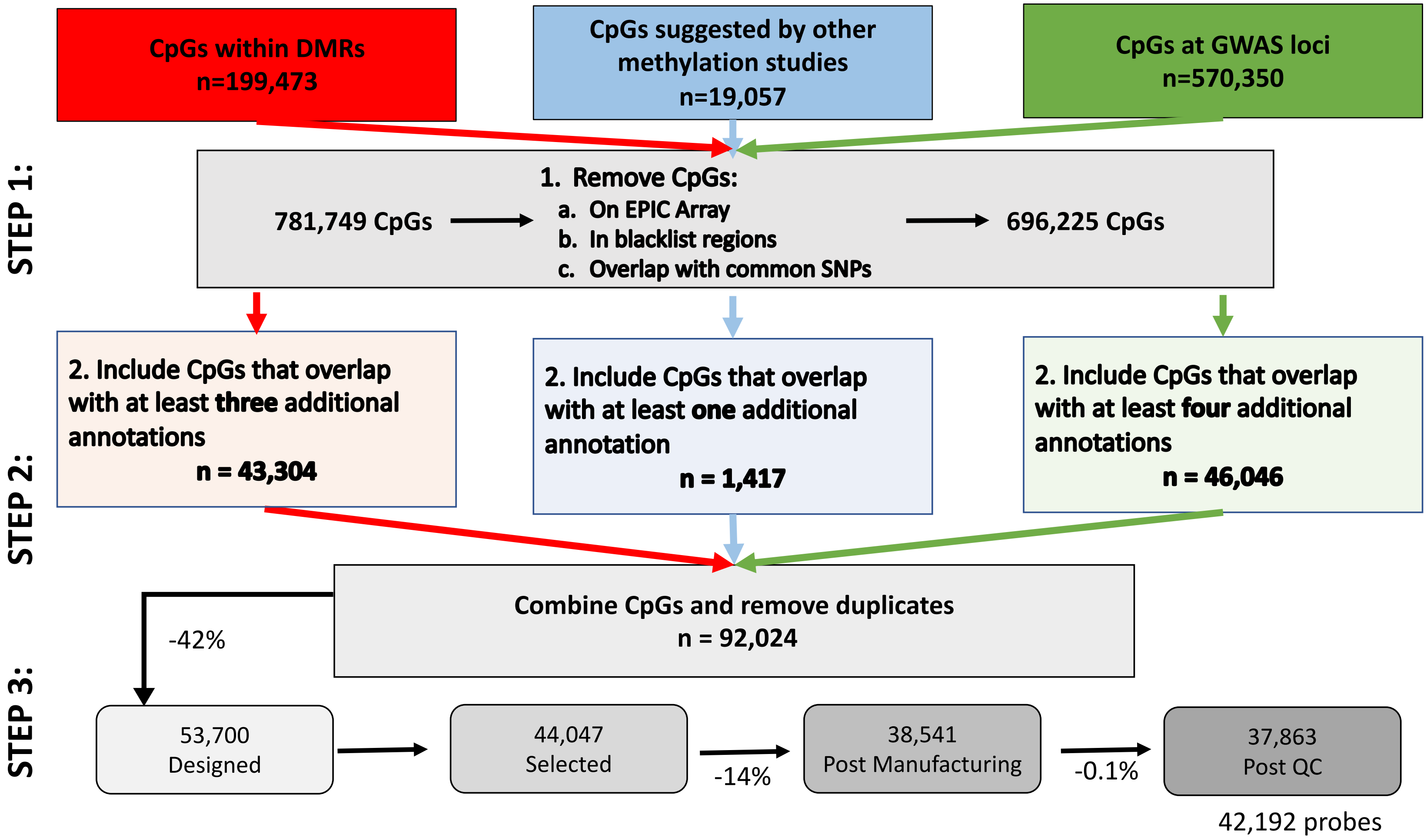

**Fig. E1. Functional genomics pipeline for identifying high-value CpGs.** The pipeline for selecting high-value CpGs involved three steps. In Step 1, we identified all CpGs with some prior evidence linking them to asthma or allergic diseases in three categories. Using

WGBS data, we identified CpG sites within asthma DMRs in ethnically diverse children (red box). Based on literature reviews, we identified CpGs associated with asthma or allergic diseases in other DNA methylation studies (blue box) and CpGs at asthma or allergic disease GWAS loci (green box). After removing duplicates, we further removed CpGs that are on the EPIC array, in blacklisted regions of the genome, or overlapped with common SNPs. In Step 2, we overlapped the CpGs with functional annotations (see Supplementary Methods) and required that CpGs in DMRs from the WGBS overlapped with at least three annotations (pink box), CpGs from prior methylation studies overlapped with one additional annotation (light blue box), and CpGs from GWAS loci overlapped with at least four additional annotations. This resulted in 92,024 "high-value" CpGs. In Step 3, we submitted the CpGs to Illumina for design and manufacture of the final set of selected CpGs. Of the 38,541 CpGs that passed manufacturing, 37,863 CpGs, corresponding to 42,192 probes, passed array QC (see Methods for additional details). Information for the CpGs on the array with respect to locations, prior evidence, and functional annotations is shown in Supplementary Dataset 1.

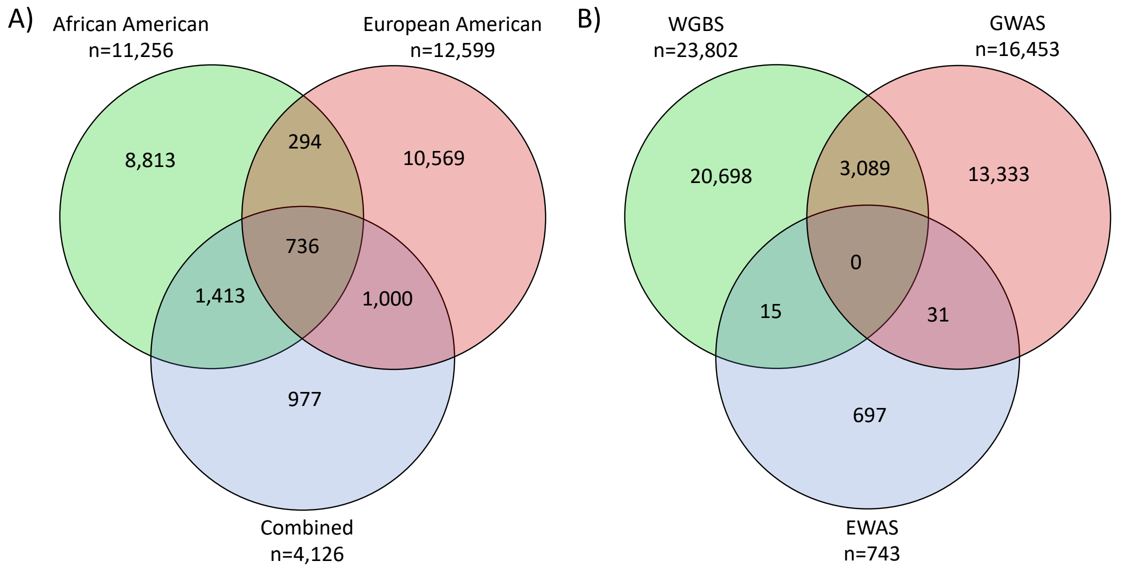

**Fig. E2. Distribution of Custom Array CpGs by primary criteria and DMR category. A**) CpGs on the Custom array proportionally represented DMR-CpGs from WGBS studies in African Americans, European Americans, and the combined samples. **B**) CpGs on the Custom array proportionally represented CpGs from the WGBS studies and previous GWAS of asthma or allergic diseases; very few previous EWAS CpGs were included on the Custom array because nearly all were on the EPIC array.

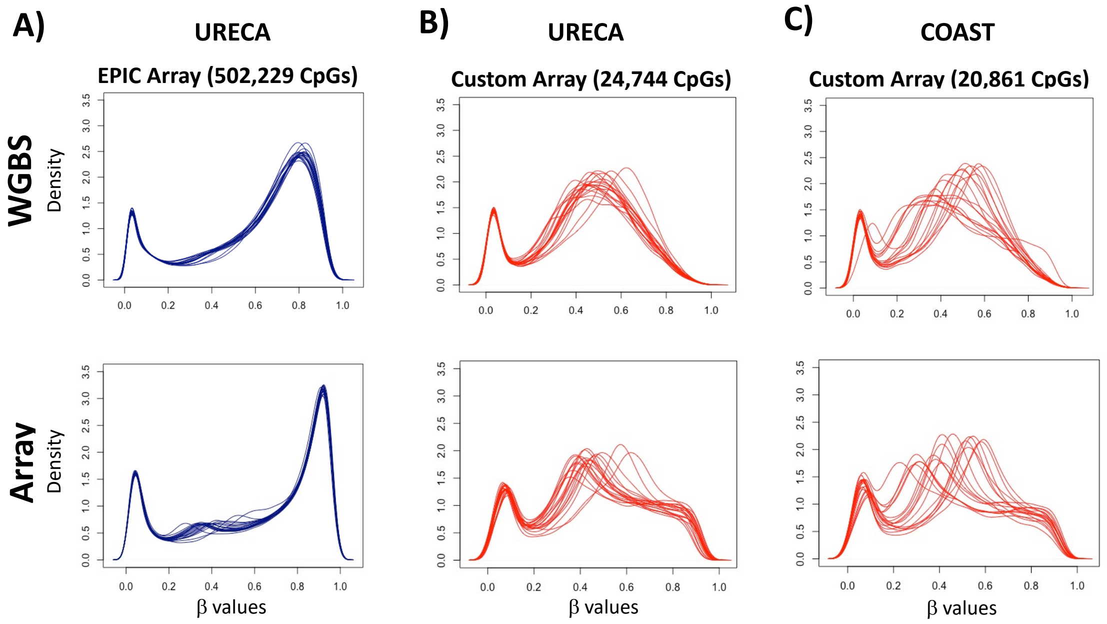

**Fig. E3. Comparison of methylation level (β value) distributions from the EPIC and Custom Arrays to the WGBS data.** Percent methylation is shown on the x-axis and density is shown on the y-axis. The number of CpGs in each comparison is shown at the top of each pair, which includes the number of sites with at least 10 mapped reads in the WGBS data. **A**) β value distribution for the WGBS data compared to the EPIC array for the 19 URECA subjects assayed using both platforms that passed QC. Spearman's rho=0.82 ($P$<2.2x$10^{-16}$). **B**) β value distribution for the WGBS data compared to the Custom array for the same 19 URECA subjects. Spearman's rho=0.83 ($P$<2.2x$10^{-16}$). **C**) β value distribution for the WGBS data compared to the Custom array for the 17 COAST samples that passed QC. Spearman's rho=0.82 ($P$<2.2x$10^{-16}$).

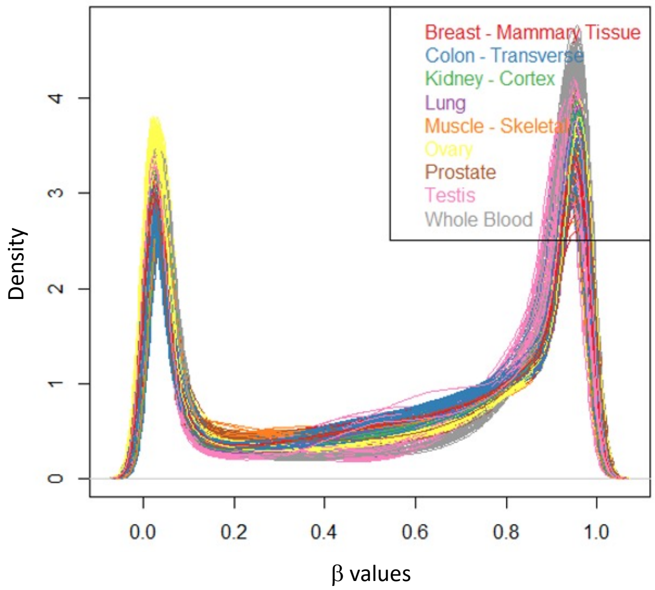

**Fig. E4. β value distribution plots from nine GTEX tissues.** Percent methylation (β value) is shown on the x-axis and density is shown on the y-axis. Data are from Oliva *et al.*, Genetic regulation of DNA methylation across tissues reveals thousands of molecular links to complex traits. *Research Square* https://doi.org/10.21203/rs.3.rs-492596/v1 (2021).

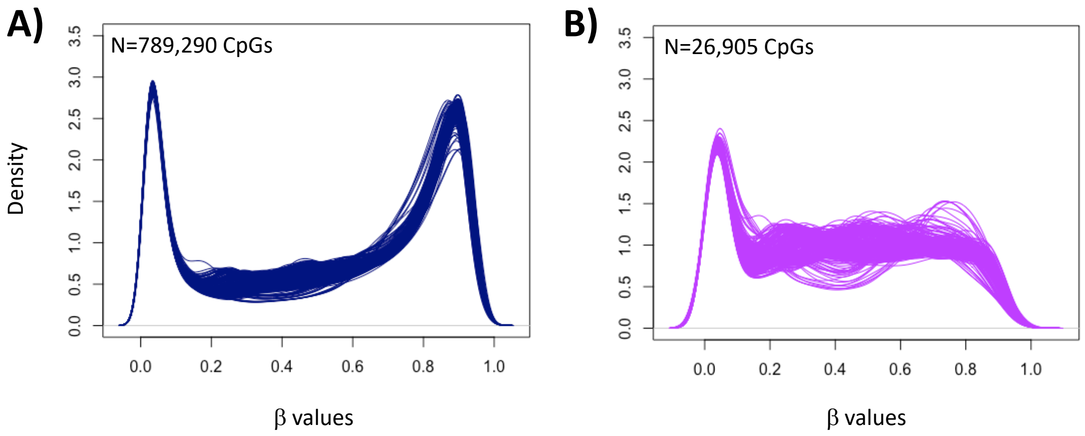

**Fig. E5. β value distributions of the high-value EPIC CpGs compared to all CpGs on the EPIC array.** To determine whether the enrichment for IM CpGs on the Custom array was attributable to the selection criteria we used for designing the array, we filtered the CpGs on the EPIC array using the same pipeline as that used for selecting CpGs for the Custom array. These are CpGs that met criteria for inclusion on the Custom array but were excluded because they were on the EPIC array. We refer to these as high-value EPIC CpGs. Of the 789,290 CpGs on the EPIC array that passed QC, 26,905 (3.4%) were designated as high-value. The β distribution of the high-value EPIC CpGs in nasal epithelial cells revealed a pattern similar to CpGs on the Custom array, with the majority (61%) having β values between 20-80% **A**) The β value distribution of the 789,290 EPIC CpGs is shown in blue. **B**) The β value distribution of the 26,905 EPIC CpGs that meet the selection criteria for inclusion on the Custom array (high-value EPIC CpGs) is shown in purple. The percent methylation (β value) is shown on the x-axis and density is shown on the y-axis.

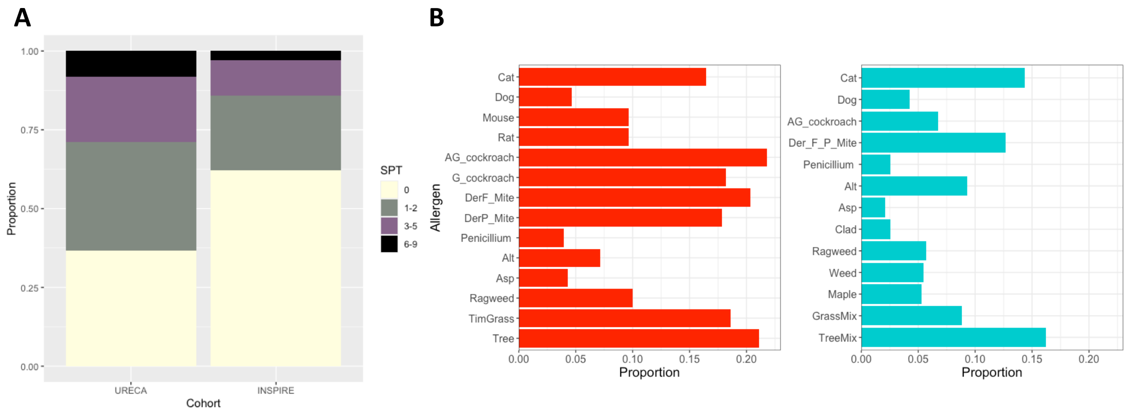

**Fig. E6. Allergic sensitization in URECA and INSPIRE. A**) Proportion of subjects with positive skin prick test (SPT) results to zero, one to two, three to five, or six to nine allergens tested in the URECA and INSPIRE cohorts. **B**) Proportion of positive skin prick tests by allergen tested in URECA and INSPIRE cohorts. The 14 allergens tested for URECA (red) include Mouse epithelia, Dog epithelia, *Dermatophagoides fainae* (mite), *Dermatophagoides pteronyssinus* (mite), Cat hair, Rat epithelia, American/German cockroach, German cockroach, *Alternaria Tenuis*, *Aspergillus* mix, Ragweed mix, Tree pollen (oak or birch), *Penicillium Notatum/Pennicillium Chrysogenum*, Timothy grass. The thirteen allergens tested for INSPIRE (turquoise) include dog, cat, *Dermatophagoides pteronyssinus* and *Dermatophagoides farinae* mix, American/German cockroach, *Penicillium Notatum/ Penicillium Chrysogenum*, *Alternaria Tenuis*, *Cladosporum Herbarum*, *Aspergillus* mix, Ragweed mix, Eastern 6 Tree mix, K-O-T Grass mix, Maple/Box Elder mix, and Weed mix.

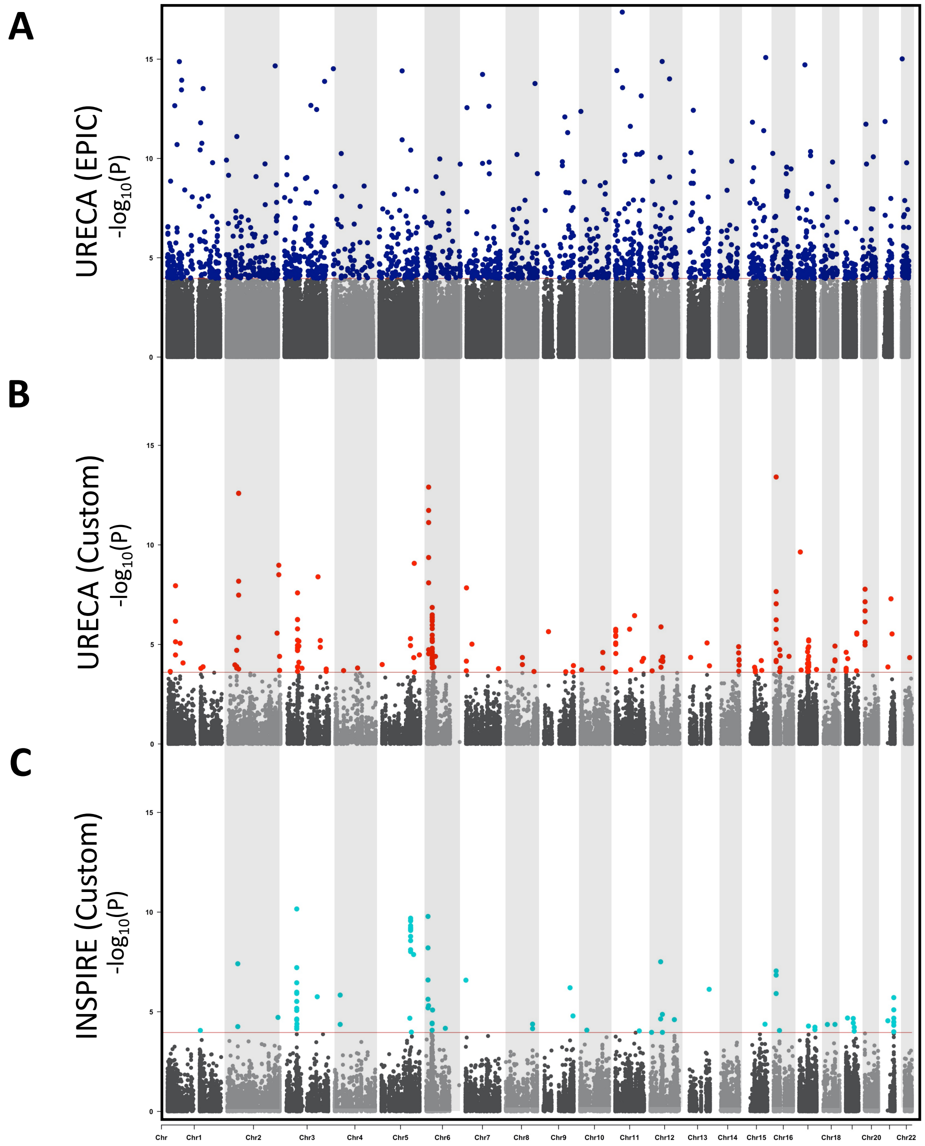

**Fig. E7. Manhattan plots illustrating EWAS results of allergic sensitization.** Chromosomes 1-22 are shown along the x-axis and -log$_{10}$ P-values are shown on the y-axis. **A)** Results in URECA (EPIC array). Significant DMCs at a q-value threshold of 0.05 are shown in blue. **B)** Results in URECA (Custom array). Significant DMCs at a q-value threshold of 0.05 are shown in red. **C)** Results in INSPIRE (Custom array). Significant DMCs at a q-value threshold of 0.05 are shown in turquoise. Plots were generated using CMplot (https:/github.com/YinLiLin/CMplot).

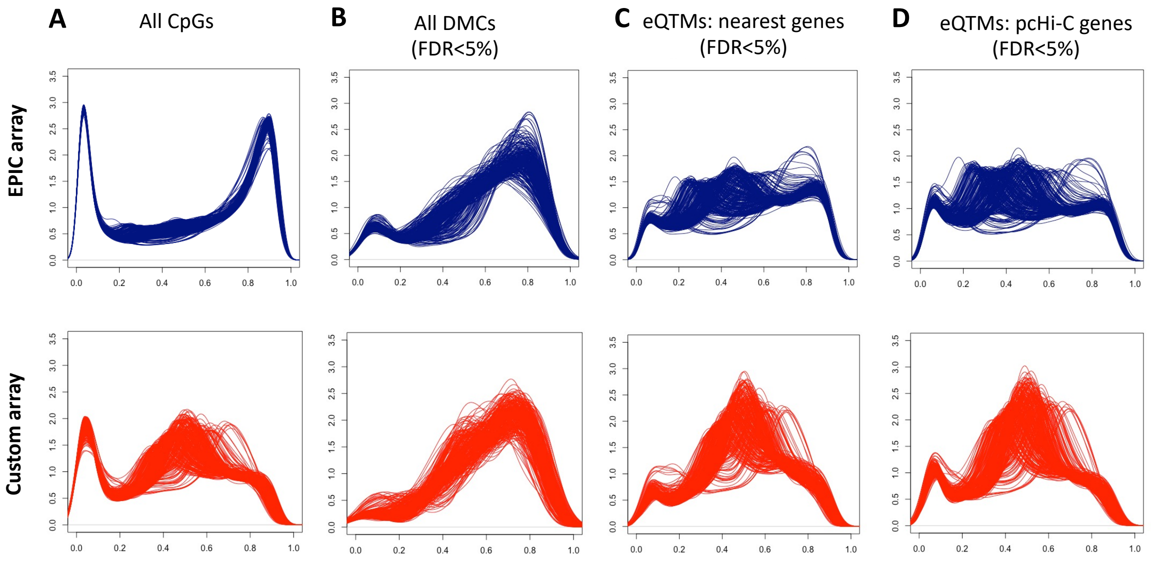

**Fig. E8. Density plots showing β value distributions for all CpGs, DMCs, eQTMs and their nearest genes, and eQTMs and their pcHi-C target genes for the EPIC and Custom arrays.** All plots show percent methylation (β value) on the x-axis and density on the y-axis. **A**) Density plots of all CpGs on the EPIC (blue; N=789,290) and Custom (red; N=37,256) arrays. **B**) Density plots of all DMCs on the EPIC (N=1,805) and Custom (N=193) arrays. **C**) Density plots of eQTMs with their nearest gene on the EPIC (N=87,193) and Custom (N=8,778) arrays. **D**) Density plots of eQTMs with their pcHi-C target genes on the EPIC (N=59,542) and Custom arrays (N=9,298) arrays.

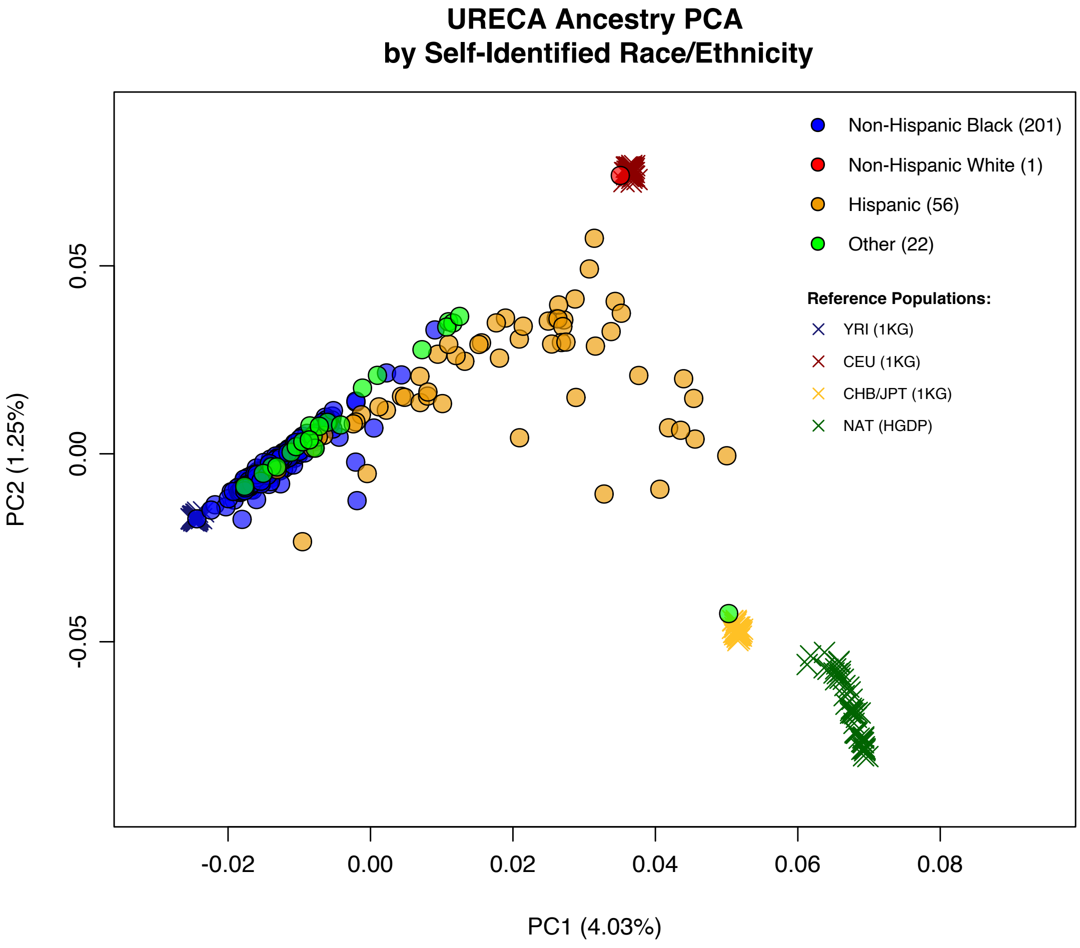

**Fig. E9. Ancestry PCA plot of self-identified race/ethnicity of 280 URECA subjects.** PC1 and PC2 are shown on the x-axis and y-axis, respectively. The proportion of variance explained is shown in parentheses.

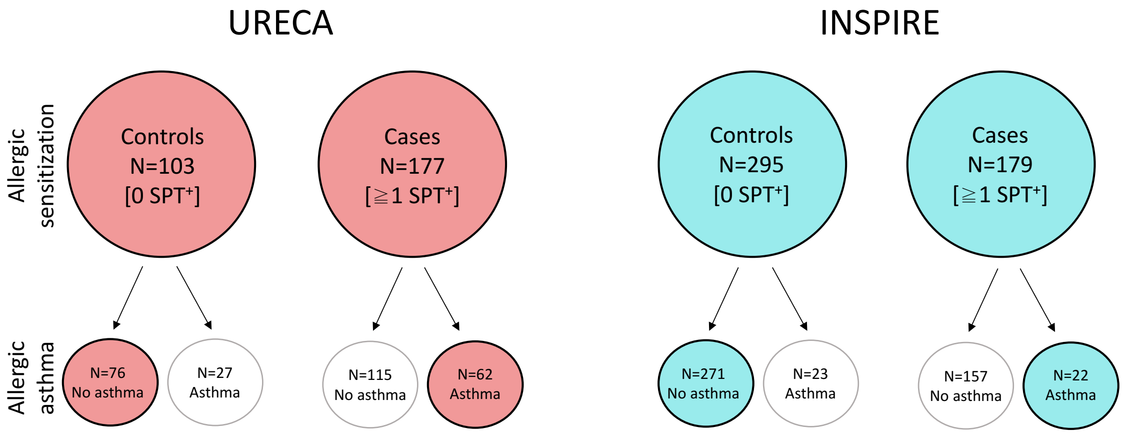

**Fig. E10. Overlap of allergic sensitization and allergic asthma cases and controls.** One INSPIRE control in the allergic sensitization (AS) EWAS was missing data for asthma diagnosis at age 6. Cases and controls for the AS EWAS are shown in larger filled circles and cases and controls for the allergic asthma EWAS are shown in the smaller filled circles.

**Fig. E11. Regional association plots for the 10 most significant DMCs in the URECA EWAS using the EPIC array.** The genomic locations and genes are shown on the x-axis; EWAS P-values are shown on the y-axis. The dashed horizontal lines show the 0.05 q-value threshold for the Custom array (red) and for the EPIC array (blue). The density of CpGs in the region is shown along the top of each plot. The most significant DMC in a region for a given EWAS (cohort and platform; see legend at lower right) is illustrated by a diamond; additional DMCs appear as circles.

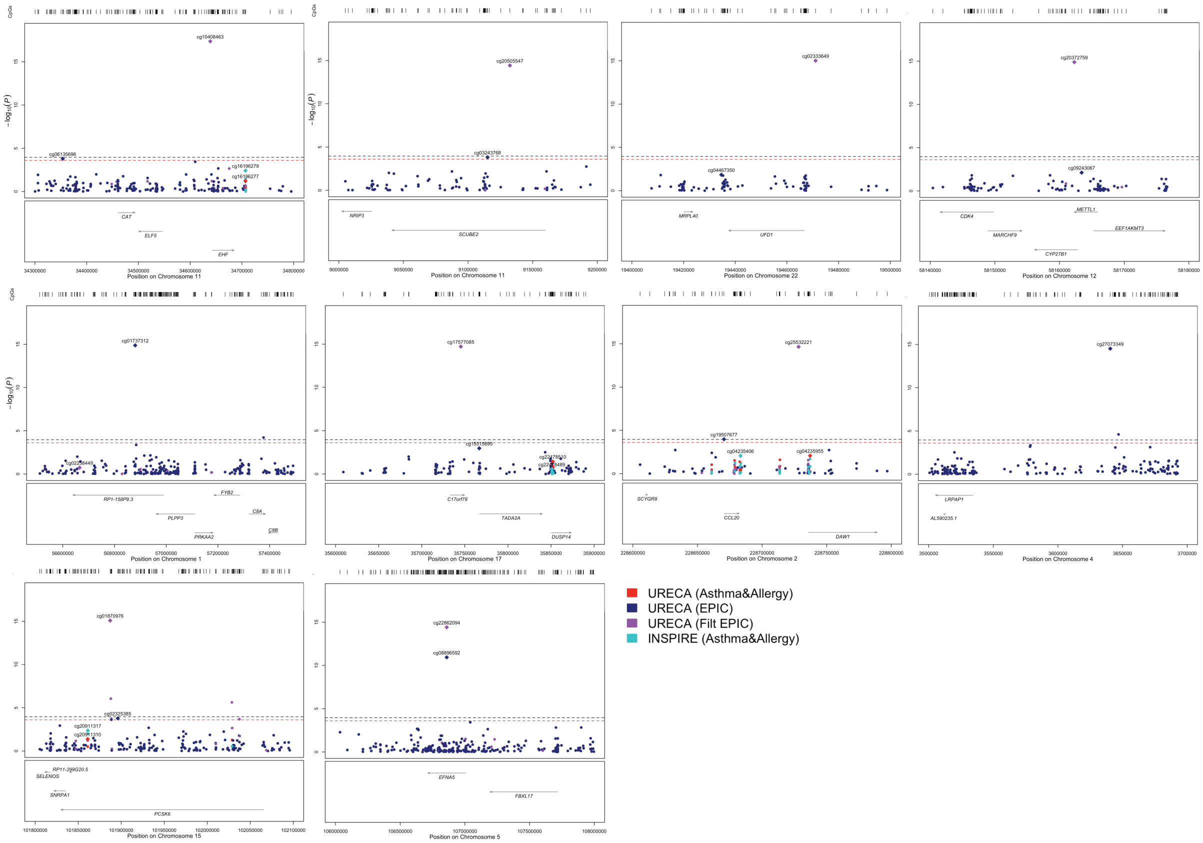

**Fig E12. Regional association plots for the 10 most significant DMCs in the URECA EWAS using the Custom array.** The genomic locations and genes are shown on the x-axis; EWAS P-values are shown on the y-axis. The dashed horizontal lines show the 0.05 q-value threshold for the Custom array (red) and for the EPIC array (blue). The density of CpGs in the region is shown along the top of each plot. The most significant DMC in a region for a given EWAS (cohort and platform; see legend upper right) is illustrated by a diamond; additional DMCs appear as circles.

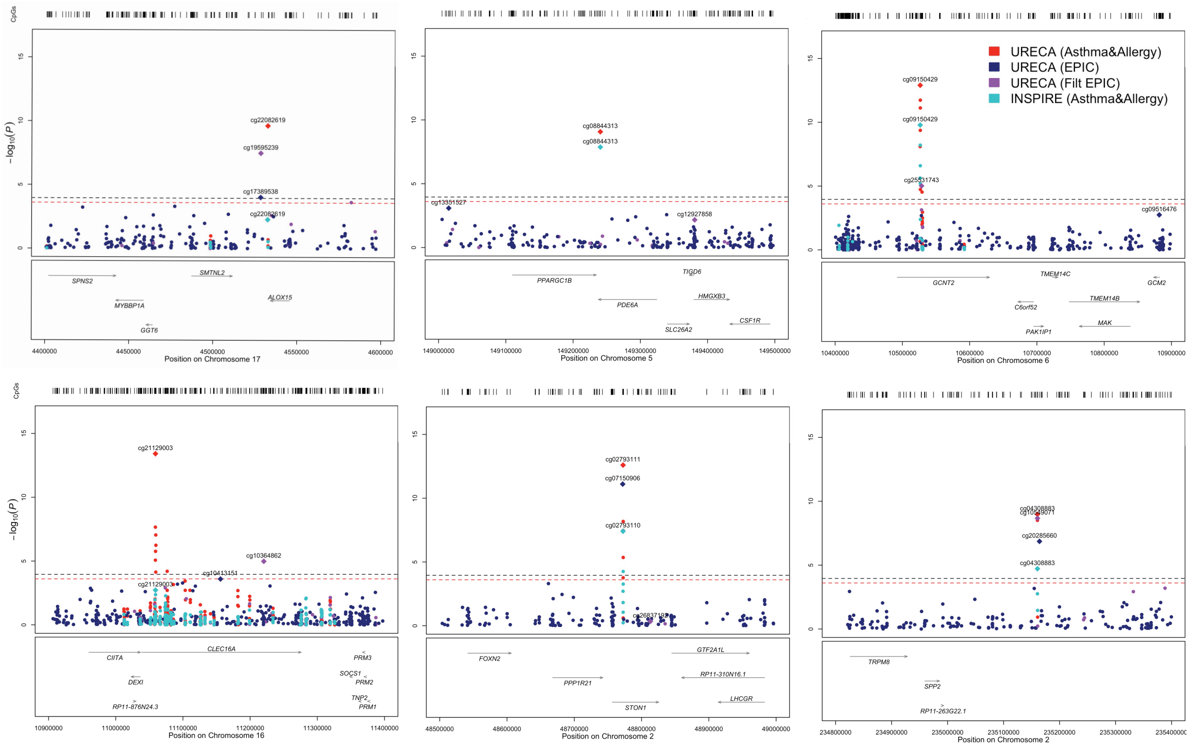

**Table E1. Description of the differentially methylated regions (DMRs) from the whole genome bisulfite sequencing study.** AA, African American; EA, European American

| Group Analyzed | # DMRs | Median Size (bp) [range] | Median # CpGs [range] | # CpGs hyper-methylated | # CpGs hypo-methylated |
|---|---|---|---|---|---|
| AA | 7,748 | 483 [6-3,828] | 10 [3-124] | 2,048 | 5,700 |
| EA | 8,972 | 437 [8-2,841] | 9 [3-144] | 1,879 | 7,093 |
| Combined | 2,585 | 513 [9-2,929] | 11 [3-163] | 498 | 2,087 |

**Table E2. EWAS studies of asthma and allergic diseases[E24-38] used for CpG selection in Step 1 (see Fig. E1).** Phenotypes are shown only for those used to select CpGs for the array.

| Illumina BeadChip | Phenotype(s) | Sample Size | Reference |
|---|---|---|---|
| **Respiratory cells** | | | |
| EPIC | Allergic rhinitis | 454 | E26 |
| 450K | Remittent asthma vs Persistent asthma or controls | 135 | E27 |
| EPIC | Asthma, FeNo, total IgE, environmental IgE, allergic asthma, bronchodilator response | 547 | E24 |
| 450K | Atopy | 483 | E25 |
| 450K | Atopic asthma | 72 | E28 |
| **Blood cells** | | | |
| 450K | Inhaled corticosteroids exposure | 215 | E31 |
| 450K | Allergic sensitization | 376 | E38 |
| 450K | Asthma | 3572 newborns<br>2862 children | E36 |
| 450K | Food allergen sensitization, allergen sensitization, atopic sensitization | 739 | E34 |
| 450K | Childhood asthma | 817 | E37 |
| 450K | Total serum IgE | 217 | E33 |
| 450K | Total serum IgE | 306 | E29 |
| 450K | Atopic sensitization and high serum IgE | 367 | E30 |
| 450k | Eczema | 366 | E35 |
| 27K | Serum IgE | 355 | E32 |

**Table E3. PCA of DNA methylation analysis and identifying batch effects**. Significance of correlations between potential confounders and PCs 1 through 5 for **A)** quantile normalized methylation data in URECA subjects (Custom), **B)** final adjusted methylation data for URECA subjects (Custom), **C)** quantile normalized methylation data in URECA subjects (EPIC), **D)** final adjusted methylation data for URECA subjects (EPIC), **E)** quantile normalized methylation data in INSPIRE subjects (Custom), and **F)** final adjusted methylation data for INSPIRE subjects (Custom). Values less than *P*=0.05 are highlighted in red.

| **A. URECA Custom Array, raw** | | | | | | | | | | | |
|---|---|---|---|---|---|---|---|---|---|---|---|
| | **PropVar** | **Plate** | **Array** | **Study_Site** | **DNA_Conc** | **Sex** | **% Ciliated** | **% Squamous** | **AncPC1** | **AncPC2** | **AncPC3** |
| PC1 | 0.386 | 0.236 | 0.016 | 0.103 | 1.37E-08 | 0.960 | 3.38E-25 | 0.008 | 0.265 | 0.198 | 0.230 |
| PC2 | 0.133 | 1.17E-90 | 4.31E-72 | 0.690 | 0.355 | 0.223 | 0.946 | 0.884 | 0.697 | 0.924 | 0.677 |
| PC3 | 0.079 | 0.002 | 0.002 | 1.72E-10 | 0.882 | 0.243 | 0.153 | 0.001 | 0.041 | 0.079 | 0.962 |
| PC4 | 0.029 | 0.246 | 0.919 | 1.54E-08 | 7.29E-07 | 0.016 | 0.896 | 0.180 | 0.150 | 0.129 | 0.361 |
| PC5 | 0.019 | 0.754 | 0.845 | 0.388 | 0.484 | 1.63E-07 | 0.243 | 0.862 | 0.186 | 0.117 | 0.893 |
| **B. URECA Custom Final. The effects of DNA concentration, plate, array, and study site were removed. Sex, % ciliated cells, ancestry PCs 1-3, and latent factors were included in the model.** | | | | | | | | | | | |
| | **PropVar** | **Plate** | **Array** | **Study_Site** | **DNA_Conc** | **Sex** | **% Ciliated** | **% Squamous** | **AncPC1** | **AncPC2** | **AncPC3** |
| PC1 | 0.389 | 0.970 | 1.000 | 0.738 | 0.839 | 0.950 | 2.15E-22 | 0.028 | 0.097 | 0.065 | 0.480 |
| PC2 | 0.089 | 0.936 | 0.999 | 0.410 | 0.436 | 0.038 | 0.231 | 0.002 | 0.130 | 0.236 | 0.817 |
| PC3 | 0.060 | 0.907 | 1.000 | 0.968 | 0.181 | 0.327 | 0.825 | 0.604 | 0.584 | 0.443 | 0.708 |
| PC4 | 0.030 | 0.982 | 1.000 | 0.296 | 0.605 | 0.059 | 0.175 | 0.362 | 0.033 | 0.151 | 0.366 |
| PC5 | 0.024 | 0.954 | 1.000 | 0.283 | 0.922 | 8.17E-07 | 0.462 | 0.614 | 0.037 | 0.030 | 0.862 |
| **C. URECA EPIC Array, raw** | | | | | | | | | | | |
| | **PropVar** | **Plate** | **Array** | **Study_Site** | **DNAConc** | **Sex** | **% Ciliated** | **% Squamous** | **AncPC1** | **AncPC2** | **AncPC3** |
| PC1 | 0.229 | 0.139 | 0.064 | 0.128 | 3.49E-08 | 0.841 | 2.62E-25 | 0.003 | 0.405 | 0.369 | 0.736 |
| PC2 | 0.064 | 0.002 | 0.002 | 3.20E-09 | 0.676 | 0.130 | 0.487 | 0.008 | 0.016 | 0.188 | 0.285 |
| PC3 | 0.021 | 0.280 | 0.176 | 0.966 | 0.826 | 0.935 | 0.247 | 0.774 | 0.089 | 0.163 | 0.773 |
| PC4 | 0.019 | 0.609 | 0.932 | 1.08E-07 | 0.010 | 0.244 | 0.172 | 0.280 | 0.025 | 0.030 | 0.130 |
| PC5 | 0.017 | 0.199 | 0.137 | 0.675 | 0.284 | 0.123 | 0.416 | 0.644 | 0.085 | 0.015 | 0.292 |

| **D. URECA EPIC Final. The effects of DNA concentration, plate, array, and study site were removed. Sex, % ciliated cells, ancestry PCs 1-3, and latent factors were included in the model.** | | | | | | | | | | | |
|---|---|---|---|---|---|---|---|---|---|---|---|
| | **PropVar** | **Plate** | **Array** | **Study_Site** | **Sex** | **DNAConc** | **% Ciliated** | **% Squamous** | **AncPC1** | **AncPC2** | **AncPC3** |
| PC1 | 0.219 | 0.925 | 0.639 | 0.727 | 0.828 | 0.645 | 8.46E-27 | 0.009 | 0.202 | 0.148 | 0.432 |
| PC2 | 0.062 | 0.773 | 0.787 | 0.489 | 0.038 | 0.940 | 0.476 | 0.028 | 0.115 | 0.202 | 0.690 |
| PC3 | 0.021 | 0.959 | 0.754 | 0.748 | 0.016 | 0.872 | 0.182 | 0.963 | 0.003 | 0.003 | 0.875 |
| PC4 | 0.016 | 0.990 | 0.997 | 0.724 | 0.002 | 0.990 | 0.883 | 0.387 | 0.513 | 0.311 | 0.915 |
| PC5 | 0.015 | 0.849 | 0.902 | 0.449 | 0.022 | 0.809 | 0.219 | 0.830 | 0.011 | 0.003 | 0.396 |

| **E. INSPIRE Custom Array, raw.** Race = parent-reported race/ethnicity. | | | | | | | | |
|---|---|---|---|---|---|---|---|---|
| | **PropVar** | **Sex** | **Race** | **DNAConc** | **Plate** | **Array** | **Age** | **Collection Yr** |
| PC1 | 0.470 | 0.081 | 0.150 | 3.53E-31 | 3.40E-22 | 1.12E-11 | 0.363 | 0.234 |
| PC2 | 0.094 | 0.963 | 0.806 | 3.66E-10 | 4.35E-11 | 3.00E-08 | 0.690 | 0.040 |
| PC3 | 0.046 | 0.896 | 0.681 | 0.188 | 0.003 | 0.276 | 0.050 | 0.759 |
| PC4 | 0.024 | 0.195 | 0.708 | 2.60E-10 | 1.92E-10 | 0.001 | 0.917 | 0.083 |
| PC5 | 0.022 | 0.147 | 0.358 | 7.09E-16 | 3.20E-98 | 2.74E-75 | 0.004 | 0.001 |

| **F. INSPIRE Custom Array Final. The effects of DNA concentration, collection year, plate, and array were removed. Sex, parent-reported race/ethnicity, DNA concentration, and latent factors were included in the model.** | | | | | | | | |
|---|---|---|---|---|---|---|---|---|
| | **PropVar** | **Sex** | **Race** | **DNAConc** | **Plate** | **Array** | **Age** | **Collection Yr** |
| PC1 | 0.445 | 0.049 | 0.450 | 0.913 | 0.999 | 1.000 | 0.776 | 0.430 |
| PC2 | 0.082 | 0.964 | 0.657 | 0.125 | 0.994 | 1.000 | 0.147 | 0.539 |
| PC3 | 0.051 | 0.673 | 0.768 | 0.064 | 0.996 | 1.000 | 0.587 | 0.891 |
| PC4 | 0.026 | 0.033 | 0.369 | 0.194 | 0.990 | 1.000 | 0.882 | 0.187 |
| PC5 | 0.014 | 0.009 | 0.043 | 0.009 | 0.929 | 1.000 | 0.063 | 0.757 |

**Table E4. Significant loci in GWAS studies of asthma and allergic disease used to define regions for CpG inclusion.** Both studies[E9, 10] were performed using data for white British subjects in the UK Biobank.

| Chr | Start[1] | End | Phenotype[2] | Study |
|---|---|---|---|---|
| 1 | 2492665 | 3175371 | Hay Fever | E10 |
| 1 | 8412989 | 9355936 | Hay Fever, Asthma | E10 |
| 1 | 12100942 | 12147311 | Hay Fever | E10 |
| 1 | 12175658 | 12175658 | Asthma | E10 |
| 1 | 25224509 | 25263997 | Hay Fever | E10 |
| 1 | 149897287 | 153166983 | Hay Fever, Asthma, Childhood onset asthma | E10, E9 |
| 1 | 154405024 | 154428283 | Hay Fever | E10 |
| 1 | 161159147 | 161187665 | Hay Fever, Asthma | E10 |
| 1 | 167198536 | 167439010 | Hay Fever, Asthma | E10 |
| 1 | 172777616 | 173171841 | Hay Fever, Asthma, Childhood onset asthma | E10, E9 |
| 1 | 198656242 | 198670555 | Asthma | E10 |
| 1 | 203058476 | 203108508 | Asthma, shared adult and childhood onset asthma | E10, E9 |
| 1 | 212858748 | 212877647 | Hay Fever | E10 |
| 2 | 8438693 | 8496062 | Hay Fever, Asthma, shared adult and childhood onset asthma | E10, E9 |
| 2 | 28623159 | 28644670 | Hay Fever | E10 |
| 2 | 61112552 | 61161095 | Hay Fever | E10 |
| 2 | 102243154 | 103277862 | Hay Fever, Asthma, shared adult and childhood onset asthma | E10, E9 |
| 2 | 112253302 | 112268892 | Hay Fever | E10 |
| 2 | 113582782 | 113689747 | Hay Fever | E10 |
| 2 | 143745800 | 143886819 | Hay Fever | E10 |
| 2 | 146111968 | 146316319 | adult onset asthma | E9 |
| 2 | 198148084 | 198954774 | Hay Fever, Asthma | E10 |
| 2 | 228625484 | 228751874 | Hay Fever, Childhood onset asthma | E10, E9 |
| 2 | 234113057 | 234115739 | Hay Fever | E10 |
| 2 | 242562010 | 242838542 | Hay Fever, Asthma, shared adult and childhood onset asthma | E10, E9 |
| 3 | 32920602 | 33146535 | Hay Fever, Asthma, shared adult and childhood onset asthma | E10, E9 |
| 3 | 50701250 | 51441307 | Asthma | E10 |
| 3 | 72394852 | 72394852 | Hay Fever | E10 |
| 3 | 112526053 | 112693753 | Hay Fever | E10 |
| 3 | 121387784 | 121728846 | Hay Fever | E10 |
| 3 | 127886957 | 128075398 | Asthma | E10 |
| 3 | 141040654 | 141158614 | Hay Fever | E10 |
| 3 | 176708724 | 176868116 | Asthma | E10 |
| 3 | 187632967 | 188457255 | Hay Fever, Asthma, Childhood onset asthma | E10, E9 |
| 3 | 196327220 | 196454053 | Hay Fever, Asthma | E10 |
| 4 | 4766265 | 4778175 | Hay Fever | E10 |
| 4 | 38599054 | 38934478 | Hay Fever, Asthma, Childhood onset asthma | E10, E9 |

| | | | | |
|---|---|---|---|---|
| 4 | 103188709 | 103590864 | Hay Fever | E10 |
| 4 | 122993500 | 124511672 | Hay Fever, Asthma, shared adult and childhood onset asthma | E10, E9 |
| 5 | 14572453 | 14701003 | Hay Fever, Asthma, shared adult and childhood onset asthma | E10, E9 |
| 5 | 35728440 | 36074412 | Hay Fever, Asthma | E10 |
| 5 | 40442869 | 40623346 | Hay Fever | E10 |
| 5 | 71695880 | 71743322 | childhood onset asthma | E9 |
| 5 | 109612633 | 110749926 | Hay Fever, Asthma, shared adult and childhood onset asthma | E10, E9 |
| 5 | 118659579 | 118739934 | Hay Fever, Asthma | E10 |
| 5 | 129917070 | 132138129 | Asthma | E10 |
| 5 | 131336105 | 132321276 | Hay Fever, Childhood onset asthma | E10, E9 |
| 5 | 133439274 | 133639311 | Hay Fever | E10 |
| 5 | 137461112 | 137605401 | Hay Fever | E10 |
| 5 | 141400028 | 141557236 | Hay Fever, Asthma | E10 |
| 5 | 156930406 | 156988798 | Asthma | E10 |
| 5 | 159896259 | 159929015 | Hay Fever, Asthma | E10 |
| 6 | 403799 | 421196 | childhood onset asthma | E9 |
| 6 | 25823774 | 33770370 | Hay Fever, Asthma, shared adult and childhood onset asthma | E10, E9 |
| 6 | 36349890 | 36380644 | Hay Fever | E10 |
| 6 | 90808352 | 91019304 | Hay Fever, Asthma, shared adult and childhood onset asthma | E10, E9 |
| 6 | 128264925 | 128294709 | Hay Fever, Asthma | E10 |
| 6 | 135624811 | 135950204 | Hay Fever, Asthma | E10 |
| 6 | 138002175 | 138262773 | Hay Fever | E10 |
| 6 | 155162163 | 155162163 | Asthma | E10 |
| 7 | 3062629 | 3174209 | Hay Fever, Asthma | E10 |
| 7 | 20371853 | 20640689 | Hay Fever, Asthma, shared adult and childhood onset asthma | E10, E9 |
| 7 | 22755688 | 22811384 | childhood onset asthma | E9 |
| 7 | 28139386 | 28259233 | Hay Fever, Asthma, shared adult and childhood onset asthma | E10, E9 |
| 7 | 76978096 | 77038945 | Hay Fever | E10 |
| 7 | 150690176 | 150690176 | Hay Fever | E10 |
| 8 | 81171813 | 81329123 | Hay Fever, Asthma, shared adult and childhood onset asthma | E10, E9 |
| 8 | 101514998 | 101519901 | Hay Fever | E10 |
| 8 | 128777719 | 128815029 | Hay Fever, Childhood onset asthma | E10, E9 |
| 9 | 5609742 | 6621066 | Hay Fever, Asthma, shared adult and childhood onset asthma | E10, E9 |
| 9 | 16715826 | 16756377 | Hay Fever | E10 |
| 9 | 101790878 | 101820718 | Hay Fever | E10 |
| 9 | 101915887 | 101989706 | Asthma | E10 |
| 9 | 117804027 | 117834931 | Hay Fever | E10 |
| 9 | 123636121 | 123707497 | Hay Fever | E10 |
| 9 | 127022266 | 127095039 | Hay Fever | E10 |
| 9 | 131455796 | 131617167 | Hay Fever, Asthma | E10 |
| 9 | 136141870 | 136155000 | Hay Fever | E10 |
| 9 | 140500443 | 140500443 | childhood onset asthma | E9 |

| | | | | |
|---|---|---|---|---|
| 10 | 5885314 | 6631223 | Hay Fever, Asthma, Childhood onset asthma | E10, E9 |
| 10 | 8095340 | 9938970 | Hay Fever, Asthma, shared adult and childhood onset asthma | E10, E9 |
| 10 | 64349979 | 64391375 | Hay Fever | E10 |
| 10 | 94342983 | 94492716 | Asthma | E10 |
| 10 | 104222963 | 104512006 | Hay Fever | E10 |
| 11 | 1110395 | 1147618 | Asthma, shared adult and childhood onset asthma | E10, E9 |
| 11 | 2237219 | 2296012 | Hay Fever | E10 |
| 11 | 36336263 | 36388519 | Hay Fever, Asthma | E10 |
| 11 | 60793330 | 60793722 | Hay Fever | E10 |
| 11 | 61543499 | 61623140 | Asthma | E10 |
| 11 | 61630104 | 61657926 | shared adult and childhood onset asthma | E9 |
| 11 | 65495211 | 65683531 | Hay Fever, Asthma, Childhood onset asthma | E10, E9 |
| 11 | 75891182 | 76377819 | Hay Fever, Asthma, shared adult and childhood onset asthma | E10, E9 |
| 11 | 95419908 | 95426984 | Hay Fever | E10 |
| 11 | 111415822 | 111647084 | Hay Fever, Asthma | E10 |
| 11 | 118550522 | 118770321 | Hay Fever, Childhood onset asthma | E10, E9 |
| 11 | 128131013 | 128200831 | Hay Fever | E10 |
| 12 | 48186563 | 48210318 | Asthma, shared adult and childhood onset asthma | E10, E9 |
| 12 | 55358844 | 57535266 | Hay Fever, Asthma, shared adult and childhood onset asthma | E10, E9 |
| 12 | 71405206 | 71585743 | Asthma, shared adult and childhood onset asthma | E10, E9 |
| 12 | 94556678 | 94604963 | Asthma | E10 |
| 12 | 111708458 | 112906415 | Hay Fever, Childhood onset asthma | E10, E9 |
| 12 | 121133037 | 121410678 | Hay Fever, Childhood onset asthma | E10, E9 |
| 12 | 122645048 | 123829116 | Hay Fever | E10 |
| 13 | 40975005 | 41502588 | Hay Fever | E10 |
| 13 | 44475398 | 44490181 | Asthma | E10 |
| 13 | 50808877 | 50811151 | Hay Fever | E10 |
| 13 | 73359692 | 74039935 | Hay Fever | E10 |
| 13 | 74039935 | 74039935 | Asthma | E10 |
| 13 | 99781378 | 100227069 | Hay Fever, Asthma, shared adult and childhood onset asthma | E10, E9 |
| 14 | 35510900 | 35864878 | Hay Fever | E10 |
| 14 | 68727506 | 68815261 | Hay Fever, Asthma, shared adult and childhood onset asthma | E10, E9 |
| 14 | 103067487 | 103387971 | Hay Fever | E10 |
| 15 | 41252202 | 41796498 | Hay Fever, Asthma | E10 |
| 15 | 61032054 | 61123862 | Hay Fever, Asthma, shared adult and childhood onset asthma | E10, E9 |
| 15 | 67371244 | 67469570 | Hay Fever, Asthma, shared adult and childhood onset asthma | E10, E9 |
| 15 | 75399102 | 75448181 | Hay Fever | E10 |
| 15 | 84556623 | 84556623 | Asthma | E10 |
| 15 | 90859095 | 91094064 | Hay Fever | E10 |

| | | | | |
|---|---|---|---|---|
| 16 | 11006011 | 11336508 | Hay Fever, Asthma, shared adult and childhood onset asthma | E10, E9 |
| 16 | 27203012 | 27417744 | Hay Fever, Asthma, shared adult and childhood onset asthma | E10, E9 |
| 16 | 50745926 | 50790158 | Asthma, shared adult and childhood onset asthma | E10, E9 |
| 17 | 4521473 | 4535314 | Hay Fever | E10 |
| 17 | 37281157 | 38897220 | Hay Fever, Asthma, Childhood onset asthma | E10, E9 |
| 17 | 40338997 | 40450012 | Asthma, Childhood onset asthma | E10, E9 |
| 17 | 43156023 | 43457886 | Hay Fever, Asthma | E10 |
| 17 | 45805811 | 45873184 | childhood onset asthma | E9 |
| 17 | 47299789 | 47481374 | Hay Fever, Asthma, shared adult and childhood onset asthma | E10, E9 |
| 18 | 48482900 | 48662349 | Hay Fever | E10 |
| 18 | 51781019 | 52366730 | Hay Fever, Asthma | E10 |
| 18 | 60005046 | 60018206 | Hay Fever, Childhood onset asthma | E10, E9 |
| 18 | 61412756 | 61627300 | Asthma, Childhood onset asthma | E10, E9 |
| 19 | 1149092 | 1171213 | childhood onset asthma | E9 |
| 19 | 33718053 | 33736279 | Hay Fever, Asthma, shared adult and childhood onset asthma | E10, E9 |
| 19 | 46219145 | 46370381 | Asthma | E10 |
| 20 | 45232161 | 45716594 | Hay Fever | E10 |
| 20 | 52168159 | 52268995 | Hay Fever | E10 |
| 20 | 62270637 | 62400021 | Hay Fever, Asthma | E10 |
| 21 | 36421331 | 36507786 | Asthma, shared adult and childhood onset asthma | E10, E9 |
| 22 | 37319425 | 37319425 | Hay Fever | E10 |
| 22 | 41798520 | 41941243 | Asthma | E10 |

### Table E5. CpGs included on the Custom array by annotation category.

| | GWAS | EWAS | WGBS | Total |
|---|---|---|---|---|
| Total | 570,350 | 19,057 | 199,473 | 781,749 |
| Total after removing excluded CpGs | | | | |
| Remove EPIC | 537,374 | 2,225 | 186,240 | 719,189 |
| Remove SNPs | 521,812 | 2,194 | 186,240 | 703,596 |
| Remove Blacklist | 514,443 | 2,193 | 186,240 | 696,225 |
| CpGs with functional annotations | | | | |
| 1 annotation | 237,717 | 507 | 51,859 | 290,341 |
| 2 annotations | 120,235 | 375 | 50,333 | 169,453 |
| 3 annotations | 67,232 | 270 | 40,744 | 106,723 |
| 4 annotations | 43,213 | 175 | 26,821 | 68,851 |
| 5 annotations | 37,404 | 85 | 13,246 | 49,594 |
| 6 annotations | 8,642 | 5 | 3,237 | 11,263 |
| Custom Array | | | | |
| Submitted | 46,046 | 1,417 | 43,304 | 92,024 |
| Designed | 23,041 | 1,105 | 34,119 | 53,700 |
| Selected | 19,666 | 904 | 27,200 | 44,047 |
| Manufacturing | 16,454 | 743 | 23,802 | 38,541 |
| Array QC | 16,453 | 743 | 23,802 | 37,863 |

**Table E6**. Nearest and pcHi-C target genes for AS DMCs on the EPIC and Custom arrays in URECA. For all DMCs on the EPIC and Custom arrays, a nearest gene and a pcHi-C target gene (if identified) were assigned (see Methods). The prior evidence category for each CpG is shown (GWAS, EWAS, DMR).

| | Nearest Genes | | | | pcHi-C genes | | | |
|---|---|---|---|---|---|---|---|---|
| | Custom | | EPIC | | Custom | | EPIC | |
| Gene name | CpG | Evidence | CpG | Evidence | CpG | Evidence | CpG | Evidence |
| ABCC5 | cg05910779_BC21 | DMR | | | | | cg01966760 | NA |
| ABCC5 | cg05910778_BC21 | DMR | | | | | cg16221425 | NA |
| ABHD14A | | | | | cg07095346_BC21 | EWAS | cg22988305 | EWAS |
| ACBD4 | | | | | cg22600575_BC21 | GWAS | cg05257097 | GWAS |
| ACBD4 | | | | | cg22600571_TC21 | GWAS | | |
| ACBD4 | | | | | cg22600572_BC21 | GWAS | | |
| ADAMTS15 | | | cg18023339 | NA | cg17084526_TC21 | DMR | cg21770200 | EWAS |
| AHNAK2 | cg20048107_TC21 | DMR | | | | | cg01177261 | DMR |
| AHNAK2 | cg20048043_BC21 | DMR | | | | | | |
| ALDOA | | | | | cg21355554_BC21 | DMR | cg04665974 | NA |
| ALDOA | | | | | cg21355555_TC21 | DMR | | |
| ALOX15 | cg22082619_BC21 | GWAS, DMR | cg19595239 | GWAS, EWAS | | | | |
| ALOX15 | | | cg17389538 | GWAS | | | | |
| ANKRD10 | | | cg27461824 | EWAS | cg19162921_BC21 | DMR | | |
| ANO1 | cg12357484_TC21 | EWAS | cg25723217 | EWAS | | | | |
| ANO1 | | | cg12849969 | NA | | | | |
| ANO1 | | | cg24308267 | NA | | | | |
| ARHGAP27 | | | cg22153994 | EWAS | cg22601529_BC21 | GWAS, DMR | cg21137244 | GWAS, EWAS |
| ARHGAP27 | | | | | cg22601528_TC21 | GWAS, DMR | | |
| ARHGEF3 | cg04967825_TC21 | DMR | cg01016119 | DMR, EWAS | cg04967816_TC21 | DMR | | |
| ARHGEF3 | cg04967816_TC21 | DMR | cg13068706 | NA | | | | |
| ARHGEF3 | cg04967826_BC21 | DMR | | | | | | |
| ARHGEF7 | | | cg20847766 | EWAS | cg19162921_BC21 | DMR | cg27461824 | EWAS |
| ATP8B1 | | | cg02550398 | NA | cg23606851_BC21 | DMR | cg18475483 | DMR |
| ATP8B1 | | | | | cg23606849_TC21 | DMR | cg02550398 | NA |
| ATP8B1 | | | | | cg23606852_BC21 | DMR | cg24331818 | DMR |
| BLOC1S6 | | | | | cg20334662_BC21 | DMR | cg13570892 | EWAS |
| BLOC1S6 | | | | | | | cg07558734 | DMR, EWAS |
| C12orf57 | | | | | cg17241870_BC21 | DMR | cg16707011 | NA |
| C14orf79 | | | cg01177261 | DMR | cg20048043_BC21 | DMR | | |
| C14orf79 | | | | | cg20048107_TC21 | DMR | | |
| C15orf48 | | | cg14661236 | NA | cg20334662_BC21 | DMR | | |
| C3orf18 | | | | | cg04902046_TC21 | DMR | cg09575750 | NA |
| C3orf18 | | | | | cg04902043_BC21 | DMR | | |
| CAPN14 | cg02630973_BC21 | DMR | cg04132353 | DMR, EWAS | | | | |
| CAPN14 | | | cg01827910 | NA | | | | |

| | | | | | | | | |
|---|---|---|---|---|---|---|---|---|
| CCNI2 | | | | | cg08524768_BC21 | GWAS | cg16525542 | GWAS |
| CCNI2 | | | | | cg08524766_BC21 | GWAS | cg23475112 | GWAS, EWAS |
| CDCA4 | | | | | cg20048043_BC21 | DMR | cg01177261 | DMR |
| CDCA4 | | | | | cg20050618_BC21 | DMR | | |
| CDCA4 | | | | | cg20048107_TC21 | DMR | | |
| CDK2 | | | | | cg17662800_BC21 | GWAS | cg17865265 | GWAS |
| CDK2 | | | | | | | cg08362736 | GWAS |
| CEP72 | | | | | cg07553021_TC21 | DMR | cg00049323 | EWAS |
| CISH | cg04902026_TC21 | DMR | cg09575750 | NA | | | | |
| CISH | cg04902028_TC21 | DMR | cg23005227 | DMR, EWAS | | | | |
| CISH | cg04902029_BC21 | DMR | cg16315329 | EWAS | | | | |
| CISH | cg04902032_TC21 | DMR | | | | | | |
| CISH | cg04902033_BC21 | DMR | | | | | | |
| CISH | cg04902034_BC21 | DMR | | | | | | |
| CISH | cg04902035_BC21 | DMR | | | | | | |
| CISH | cg04902037_BC21 | DMR | | | | | | |
| CISH | cg04902041_BC21 | DMR | | | | | | |
| CISH | cg04902043_BC21 | DMR | | | | | | |
| CISH | cg04902046_TC21 | DMR | | | | | | |
| CLEC16A | cg21129219_BC21 | GWAS, DMR | cg10364862 | GWAS, EWAS | | | | |
| CLEC16A | cg21129001_TC21 | GWAS | | | | | | |
| CLEC16A | cg21129004_TC21 | GWAS | | | | | | |
| CLEC16A | cg21129010_BC21 | GWAS | | | | | | |
| CLEC16A | cg21129002_BC21 | GWAS | | | | | | |
| CLEC16A | cg21129003_BC21 | GWAS | | | | | | |
| CLEC16A | cg21129011_BC21 | GWAS | | | | | | |
| CLEC16A | cg21129012_TC21 | GWAS | | | | | | |
| CLMP | cg17016940_BC21 | DMR | cg01941390 | NA | | | | |
| CLMP | | | cg16343910 | NA | | | | |
| CLMP | | | cg25379149 | NA | | | | |
| COASY | | | | | cg22547525_TC21 | GWAS | cg06848514 | NA |
| COASY | | | | | | | cg02691389 | EWAS |
| COASY | | | | | | | cg17177779 | NA |
| CTSC | cg16735276_BC21 | DMR | cg08522340 | DMR, EWAS | | | | |
| CTSC | | | cg16118839 | DMR, EWAS | | | | |
| CTSC | | | cg09706192 | DMR | | | | |
| CYB561D2 | | | | | cg04902046_TC21 | DMR | cg15152301 | NA |
| CYB561D2 | | | | | cg04902043_BC21 | DMR | cg09575750 | NA |
| CYB561D2 | | | | | | | | |
| CYB561D2 | | | | | | | | |
| CYP11A1 | cg22186216_TC21 | EWAS | cg25788983 | NA | | | | |
| DCAKD | | | | | cg22596680_TC21 | GWAS | cg00146864 | GWAS, DMR, EWAS |
| DCAKD | | | | | cg22601159_TC21 | GWAS | cg00897875 | GWAS, DMR, EWAS |
| DCAKD | | | | | cg22601346_TC21 | GWAS, DMR | cg20864568 | GWAS, DMR, EWAS |
| DCAKD | | | | | cg22601347_TC21 | GWAS, DMR | cg23315838 | NA |
| DEF6 | | | | | cg09420574_TC21 | DMR | cg09649521 | NA |

| | | | | | | | | |
|---|---|---|---|---|---|---|---|---|
| EFTUD2 | | | cg24508472 | NA | cg22596680_TC21 | GWAS | | |
| EIF2B5 | | | cg20935483 | NA | cg05910779_BC21 | DMR | cg01966760 | NA |
| EIF2B5 | | | cg14141843 | EWAS | cg05910778_BC21 | DMR | cg16221425 | NA |
| ERBB2 | | | cg143M77681 | GWAS, EWAS | cg22513183_BC21 | GWAS | | |
| EXOC3 | cg07553021_TC21 | DMR | cg00049323 | EWAS | cg07553021_TC21 | DMR | cg00049323 | EWAS |
| FMNL1 | | | | | cg22601346_TC21 | GWAS, DMR | cg00146864 | GWAS, DMR, EWAS |
| FMNL1 | | | | | cg22601347_TC21 | GWAS, DMR | cg00897875 | GWAS, DMR, EWAS |
| FMNL1 | | | | | cg22601529_BC21 | GWAS, DMR | cg20864568 | GWAS, DMR, EWAS |
| FMNL1 | | | | | cg22601159_TC21 | GWAS | cg21137244 | GWAS, EWAS |
| FMNL1 | | | | | cg22596680_TC21 | GWAS | | |
| FMNL1 | | | | | cg22601528_TC21 | GWAS, DMR | | |
| FOXP1 | cg05089320_TC21 | DMR | cg06262288 | NA | | | | |
| GCNT2 | cg09150434_TC21 | DMR | cg25531743 | DMR | | | | |
| GCNT2 | cg09150474_TC21 | DMR | | | | | | |
| GCNT2 | cg09150429_TC21 | DMR | | | | | | |
| GCNT2 | cg09150432_TC21 | DMR | | | | | | |
| GCNT2 | cg09150433_BC21 | DMR | | | | | | |
| GCNT2 | cg09150431_BC21 | DMR | | | | | | |
| GCNT2 | cg09150428_TC21 | DMR | | | | | | |
| GDF9 | | | | | cg08524766_BC21 | GWAS | cg23475112 | GWAS, EWAS |
| GDF9 | | | | | cg08524768_BC21 | GWAS | | |
| GNAI2 | | | | | cg04902043_BC21 | DMR | cg14433598 | NA |
| GNAI2 | | | | | cg04902046_TC21 | DMR | cg09579833 | EWAS |
| GNG12 | cg00902711_TC21 | DMR | cg19277299 | NA | | | cg19277299 | NA |
| GRB7 | | | | | cg22513183_BC21 | GWAS | cg14377681 | EWAS, GWAS |
| H6PD | | | cg09869882 | NA | cg00185221_TC21 | GWAS | | |
| HEXIM1 | | | | | cg22596680_TC21 | GWAS | cg00146864 | GWAS, DMR, EWAS |
| HEXIM1 | | | | | cg22601346_TC21 | GWAS, DMR | cg00897875 | GWAS, DMR, EWAS |
| HEXIM1 | | | | | cg22601347_TC21 | GWAS, DMR | cg20864568 | GWAS, DMR, EWAS |
| HEXIM1 | | | | | cg22601528_TC21 | GWAS, DMR | cg21137244 | GWAS, EWAS |
| HEXIM1 | | | | | cg22601529_BC21 | GWAS, DMR | | |
| HYAL1 | | | | | cg04902046_TC21 | DMR | cg09575750 | NA |
| HYAL1 | | | | | cg04902043_BC21 | DMR | | |
| INO80 | | | | | cg20291832_BC21 | GWAS | cg25454569 | NA |
| IRF1 | | | | | cg08524766_BC21 | GWAS | cg23475112 | GWAS, EWAS |
| IRF1 | | | | | cg08524768_BC21 | GWAS | | |
| ITIH4 | cg07095346_BC21 | EWAS | | | | | cg09469170 | EWAS |
| JAZF1 | cg10813825_TC21 | GWAS | cg04266607 | GWAS | | | | |
| JAZF1 | | | cg06607889 | NA | | | | |
| KDM8 | | | | | cg21317192_TC21 | GWAS | cg07057349 | NA |

| | | | | | | | | |
|---|---|---|---|---|---|---|---|---|
| KIF18B | | | | | cg22601346_TC21 | GWAS, DMR | cg00146864 | GWAS, DMR, EWAS |
| KIF18B | | | | | cg22596680_TC21 | GWAS | cg00897875 | GWAS, DMR, EWAS |
| KIF18B | | | | | cg22601347_TC21 | GWAS, DMR | cg20864568 | GWAS, DMR, EWAS |
| LRRFIP1 | cg04356823_TC21 | DMR | cg02797113 | DMR, EWAS | | | cg16630940 | EWAS |
| LRRFIP1 | cg04356831_BC21 | DMR | cg16630940 | EWAS | | | | |
| LRRFIP1 | cg04356833_BC21 | DMR | | | | | | |
| LRRFIP1 | | | | | | | | |
| LRRFIP1 | | | | | | | | |
| LRRFIP1 | | | | | | | | |
| LY96 | cg12730697_BC21 | DMR | cg22007804 | NA | | | cg22007804 | NA |
| LY96 | cg12730695_TC21 | DMR | | | | | | |
| MAD1L1 | cg10556822_BC21 | DMR | cg18752987 | NA | cg10556822_BC21 | DMR | | |
| MANF | | | | | cg04902046_TC21 | DMR | cg23005227 | DMR, EWAS |
| MANF | | | | | cg04902026_TC21 | DMR | | |
| MANF | | | | | cg04902033_BC21 | DMR | | |
| MANF | | | | | cg04902034_BC21 | DMR | | |
| MANF | | | | | cg04902037_BC21 | DMR | | |
| MANF | | | | | cg04902041_BC21 | DMR | | |
| MANF | | | | | cg04902035_BC21 | DMR | | |
| MANF | | | | | cg04902028_TC21 | DMR | | |
| MANF | | | | | cg04902032_TC21 | DMR | | |
| MANF | | | | | cg04902029_BC21 | DMR | | |
| MANF | | | | | cg04902043_BC21 | DMR | | |
| MAP3K14 | cg22600571_TC21 | GWAS | cg00146864 | GWAS, DMR, EWAS | cg22601159_TC21 | GWAS | | |
| MAP3K14 | cg22600572_BC21 | GWAS | cg00897875 | GWAS, DMR, EWAS | | | | |
| MAP3K14 | cg22600575_BC21 | GWAS | cg05257097 | GWAS | | | | |
| MAP3K14 | cg22601159_TC21 | GWAS | cg16022555 | GWAS | | | | |
| MAP3K14 | cg22601346_TC21 | GWAS, DMR | cg20864568 | GWAS, DMR, EWAS | | | | |
| MAP3K14 | cg22601347_TC21 | GWAS, DMR | cg21137244 | GWAS, EWAS | | | | |
| MAP3K14 | cg22601528_TC21 | GWAS, DMR | | | | | | |
| MAP3K14 | cg22601529_BC21 | GWAS, DMR | | | | | | |
| MMP19 | cg17662800_BC21 | GWAS | cg17865265 | GWAS | | | | |
| MMP19 | | | cg08362736 | GWAS | | | | |
| MRPS6 | cg25807822_BC21 | DMR | cg25011666 | NA | | | | |
| MRPS6 | | | cg21291385 | EWAS | | | | |
| MYC | cg13109596_TC21 | GWAS | cg08349436 | NA | | | | |
| MYC | | | cg03691530 | EWAS | | | | |
| MYC | | | cg21975232 | NA | | | | |
| MYC | | | cg26169156 | NA | | | | |
| NAT6 | | | | | cg04902029_BC21 | DMR | cg23005227 | DMR, EWAS |
| NAT6 | | | | | cg04902046_TC21 | DMR | | |
| NAT6 | | | | | cg04902034_BC21 | DMR | | |
| NAT6 | | | | | cg04902028_TC21 | DMR | | |
| NAT6 | | | | | cg04902032_TC21 | DMR | | |
| NAT6 | | | | | cg04902043_BC21 | DMR | | |
| NAT6 | | | | | cg04902033_BC21 | DMR | | |

| | | | | | | | | |
|---|---|---|---|---|---|---|---|---|
| NAT6 | | | | | cg04902037_BC21 | DMR | | |
| NAT6 | | | | | cg04902035_BC21 | DMR | | |
| NAT6 | | | | | cg04902026_TC21 | DMR | | |
| NAT6 | | | | | cg04902041_BC21 | DMR | | |
| NEDD4L | cg23606849_TC21 | DMR | cg18475483 | DMR | | | | |
| NEDD4L | cg23606851_BC21 | DMR | cg24331818 | DMR | | | | |
| NEDD4L | cg23606852_BC21 | DMR | | | | | | |
| NR1D1 | | | | | cg22515799_TC21 | GWAS, DMR | cg13762512 | GWAS |
| NR1D1 | | | | | cg22515797_BC21 | GWAS | | |
| NRIP1 | cg21021629_BC21 | EWAS | cg00712106 | EWAS | | | | |
| OIP5 | | | | | cg20291832_BC21 | GWAS | cg25454569 | NA |
| P4HA2 | | | cg16476284 | NA | cg08524766_BC21 | GWAS | cg23475112 | GWAS, EWAS |
| P4HA2 | | | | | cg08524768_BC21 | GWAS | | |
| PARL | | | cg01966760 | NA | cg05910779_BC21 | DMR | | |
| PARL | | | cg16221425 | NA | cg05910778_BC21 | DMR | | |
| PELP1 | | | | | cg22082619_BC21 | GWAS, DMR | cg17389538 | GWAS |
| PELP1 | | | | | | | cg19595239 | GWAS, EWAS |
| PFKFB3 | | | cg22750548 | GWAS | cg14607001_TC21 | GWAS | cg04808066 | NA |
| PLCD3 | | | | | cg22601346_TC21 | GWAS, DMR | cg20864568 | GWAS, DMR, EWAS |
| PLCD3 | | | | | cg22596680_TC21 | GWAS | cg00897875 | GWAS, DMR, EWAS |
| PLCD3 | | | | | cg22601347_TC21 | GWAS, DMR | | |
| PRDM10 | cg17084526_TC21 | DMR | cg25007761 | NA | cg17084526_TC21 | DMR | cg25007761 | NA |
| PRDM10 | | | cg06182390 | NA | | | | |
| PRKAG2 | cg11918680_TC21 | DMR | cg26405880 | NA | cg11918680_TC21 | DMR | cg09932376 | NA |
| PRKAG2 | | | cg04578183 | NA | | | | |
| PRKAG2 | | | cg09932376 | NA | | | | |
| RAB38 | | | cg00167102 | NA | cg16735276_BC21 | DMR | cg09706192 | DMR |
| RAB38 | | | | | | | cg08522340 | DMR, EWAS |
| RAB38 | | | | | | | cg16118839 | DMR, EWAS |
| RAD50 | | | | | cg08524768_BC21 | GWAS | cg23475112 | GWAS, EWAS |
| RAD50 | | | | | cg08524766_BC21 | GWAS | | |
| RASSF1 | | | | | cg04902026_TC21 | DMR | cg09575750 | NA |
| RASSF1 | | | | | cg04902028_TC21 | DMR | cg23005227 | DMR, EWAS |
| RASSF1 | | | | | cg04902029_BC21 | DMR | | |
| RASSF1 | | | | | cg04902032_TC21 | DMR | | |
| RASSF1 | | | | | cg04902033_BC21 | DMR | | |
| RASSF1 | | | | | cg04902034_BC21 | DMR | | |
| RASSF1 | | | | | cg04902035_BC21 | DMR | | |
| RASSF1 | | | | | cg04902037_BC21 | DMR | | |
| RMI2 | | | | | cg21129011_BC21 | GWAS | cg10364862 | GWAS, EWAS |
| RMI2 | | | | | cg21129012_TC21 | GWAS | | |
| RMI2 | | | | | cg21129003_BC21 | GWAS | | |
| RMI2 | | | | | cg21129010_BC21 | GWAS | | |
| RMI2 | | | | | cg21129004_TC21 | GWAS | | |

| | | | | | | | | |
|---|---|---|---|---|---|---|---|---|
| RMI2 | | | | | cg21129002_BC21 | GWAS | | |
| RMI2 | | | | | cg21129001_TC21 | GWAS | | |
| RPL7 | | | | | cg12730695_TC21 | DMR | cg18305583 | NA |
| RPL7 | | | | | cg12730697_BC21 | DMR | | |
| S100PBP | | | cg02962744 | NA | cg00549862_BC21 | DMR | | |
| S100PBP | | | | | cg00549865_BC21 | DMR | | |
| S100PBP | | | | | cg00549864_TC21 | DMR | | |
| SEPT8 | | | cg16525542 | GWAS | cg08524766_BC21 | GWAS | cg23475112 | GWAS, EWAS |
| SEPT8 | | | | | cg08524768_BC21 | GWAS | | |
| SHROOM1 | | | | | cg08524766_BC21 | GWAS | cg23475112 | GWAS, EWAS |
| SHROOM1 | | | | | cg08524768_BC21 | GWAS | | |
| SLC12A7 | | | | | cg07553021_TC21 | DMR | cg00049323 | EWAS |
| SLC22A5 | cg08524768_BC21 | GWAS | cg23475112 | GWAS, EWAS | | | cg16476284 | GWAS |
| SLC22A5 | cg08524766_BC21 | GWAS | | | | | | |
| SLC25A39 | | | cg19481596 | NA | cg22547036_TC21 | GWAS | cg00897875 | GWAS, DMR, EWAS |
| SLC25A39 | | | | | cg22547038_BC21 | GWAS | cg20864568 | GWAS, DMR, EWAS |
| SLC25A39 | | | | | cg22601347_TC21 | GWAS, DMR | | |
| SLC25A39 | | | | | cg22601346_TC21 | GWAS, DMR | | |
| SLC48A1 | | | | | cg17561368_TC21 | DMR | cg15635287 | NA |
| SLC48A1 | | | | | cg17561366_TC21 | DMR | cg26115531 | EWAS |
| SLC48A1 | | | | | cg17561362_TC21 | DMR | | |
| SNX8 | cg10562002_BC21 | DMR | cg06047184 | DMR | | | | |
| SOCS1 | | | | | cg21129001_TC21 | GWAS | cg10364862 | GWAS, EWAS |
| SOCS1 | | | | | cg21129010_BC21 | GWAS | | |
| SOCS1 | | | | | cg21129004_TC21 | GWAS | | |
| SOCS1 | | | | | cg21129003_BC21 | GWAS | | |
| SOCS1 | | | | | cg21129219_BC21 | GWAS, DMR | | |
| SOCS1 | | | | | cg21129002_BC21 | GWAS | | |
| SOCS1 | | | | | cg21129011_BC21 | GWAS | | |
| SOCS1 | | | | | cg21129012_TC21 | GWAS | | |
| SPATA32 | | | | | cg22601346_TC21 | GWAS, DMR | cg00897875 | GWAS, DMR, EWAS |
| SPATA32 | | | | | cg22601347_TC21 | GWAS, DMR | cg20864568 | GWAS, DMR, EWAS |
| SPATA32 | | | | | | | cg00146864 | GWAS, DMR, EWAS |
| SSBP3 | cg00792117_TC21 | DMR | cg19682405 | NA | | | | |
| SSR3 | cg05713868_BC21 | DMR | | | | | cg20877312 | NA |
| SSR3 | cg05713870_BC21 | DMR | | | | | | |
| STAT3 | | | | | cg22547525_TC21 | GWAS | cg02691389 | EWAS |
| STAT3 | | | | | cg22547036_TC21 | GWAS | cg17177779 | NA |
| STAT3 | | | | | cg22547038_BC21 | GWAS | cg06848514 | NA |
| STON1 | cg02793110_BC21 | DMR | cg03390090 | NA | | | | |
| STON1 | cg02793111_BC21 | DMR | cg07150906 | DMR | | | | |
| STON1 | cg02793115_BC21 | DMR | cg10463553 | NA | | | | |
| STON1 | cg02793116_TC21 | DMR | cg23971565 | NA | | | | |

| | | | | | | | | |
|---|---|---|---|---|---|---|---|---|
| STON1 | cg02793118_TC21 | DMR | | | | | | |
| STON1 | cg02793119_TC21 | DMR | | | | | | |
| SYNPO | | | cg25162888 | EWAS | cg08706842_TC21 | DMR | | |
| SYNPO | | | cg06675531 | EWAS | | | | |
| TAOK2 | | | | | cg21355555_TC21 | DMR | cg04665974 | NA |
| TAOK2 | | | | | cg21355554_BC21 | DMR | | |
| TMEM14C | | | | | cg09150474_TC21 | DMR | cg25531743 | DMR |
| TMEM54 | | | | | cg00549865_BC21 | DMR | cg02962744 | NA |
| TMEM54 | | | | | cg00549864_TC21 | DMR | | |
| TMEM54 | | | | | cg00549861_TC21 | DMR | | |
| TMEM54 | | | | | cg00549862_BC21 | DMR | | |
| TPPP | | | | | cg07553021_TC21 | DMR | cg00049323 | EWAS |
| TRERF1 | cg09496657_BC21 | DMR | cg19406053 | NA | | | | |
| TRIM69 | cg20334662_BC21 | DMR | cg09359575 | NA | | | | |
| TRPM8 | cg04308881_TC21 | DMR | cg10549071 | DMR, EWAS | cg04308881_TC21 | DMR | cg10549071 | DMR, EWAS |
| TRPM8 | cg04308883_BC21 | DMR | cg20285660 | NA | cg04308883_BC21 | DMR | | |
| TRPM8 | | | | | | | | |
| TRPM8 | | | | | | | | |
| TRPM8 | | | | | | | | |
| TRPM8 | | | | | | | | |
| TRPM8 | | | | | | | | |
| TRPM8 | | | | | | | | |
| WIBG | | | | | cg17662800_BC21 | GWAS | cg08362736 | GWAS |
| WIBG | | | | | | | cg17865265 | GWAS |
| ZMYND10 | | | | | cg04902026_TC21 | DMR | cg09575750 | NA |
| ZMYND10 | | | | | cg04902028_TC21 | DMR | cg09579833 | EWAS |
| ZMYND10 | | | | | cg04902029_BC21 | DMR | cg23005227 | DMR, EWAS |
| ZMYND10 | | | | | cg04902032_TC21 | DMR | | |
| ZMYND10 | | | | | cg04902033_BC21 | DMR | | |
| ZMYND10 | | | | | cg04902034_BC21 | DMR | | |
| ZMYND10 | | | | | cg04902035_BC21 | DMR | | |
| ZMYND10 | | | | | cg04902037_BC21 | DMR | | |
| ZMYND10 | | | | | cg04902041_BC21 | DMR | | |
| ZMYND10 | | | | | cg04902043_BC21 | DMR | | |
| ZMYND10 | | | | | cg04902046_TC21 | DMR | | |

**Table E7. Results of KEGG pathway analysis on genes nearest DMCs and pcHi-C target genes on the Custom array using iPathway Guide.** The list of genes (N=318) was submitted to iPathway Guide using the total list of genes nearest CpGs on the Custom and EPIC arrays combined (N=14,049) as background. The "countAll" column lists the number of genes in the background list that fall into the pathways shown. The corresponding list of genes for the EPIC array did not reveal any enriched pathways.

| Pathway | countDE | countAll | fdr |
|---|---|---|---|
| Prolactin signaling pathway | 9 | 62 | 0.011 |
| Th1 and Th2 cell differentiation | 6 | 84 | 0.022 |
| Viral carcinogenesis | 13 | 164 | 0.022 |
| Adipocytokine signaling pathway | 8 | 55 | 0.022 |
| JAK-STAT signaling pathway | 11 | 119 | 0.023 |
| Acute myeloid leukemia | 8 | 61 | 0.024 |
| Human T-cell leukemia virus 1 infection | 15 | 203 | 0.024 |
| PI3K-Akt signaling pathway | 17 | 279 | 0.024 |
| Th17 cell differentiation | 8 | 97 | 0.024 |
| Small cell lung cancer | 9 | 85 | 0.029 |
| Non-small cell lung cancer | 7 | 66 | 0.029 |
| Hippo signaling pathway - multiple species | 1 | 27 | 0.042 |
| Epstein-Barr virus infection | 12 | 175 | 0.042 |
| Viral protein interaction with cytokine and cytokine receptor | 2 | 71 | 0.042 |
| Pathways in cancer | 22 | 449 | 0.044 |
| Alcoholic liver disease | 7 | 117 | 0.045 |
| PPAR signaling pathway | 1 | 56 | 0.049 |

**Table E8. Enrichment of eQTMs among all CpGs and among DMCs on the Custom and EPIC arrays.**

| | # of CpGs | Percent eQTMs | |
|---|---|---|---|
| | | **Nearest Gene** | **pcHi-C Target Gene** |
| EPIC | 789,290 | 11% | 9% |
| Custom | 37,256 | 23% | 16% |
| *P*-value for difference (FET) | - | $<2.2 \times 10^{-16}$ | $<2.2 \times 10^{-16}$ |
| | **# of DMCs** | **Percent eQTMs** | |
| | | **Nearest Gene** | **pcHi-C Target Gene** |
| EPIC | 1,805 | 20% | 15% |
| Custom | 193 | 35% | 22% |
| *P*-value for difference (FET) | - | 0.0019 | 0.0082 |

**Table E9. Annotation information for three selected loci.** For each locus, the number of DMCs (URECA and INSPIRE), eQTMs for the nearest gene and pcHi-C target gene, genic location, primary inclusion criteria (GWAS, EWAS, and/or DMR), and functional annotation category are shown for each platform (Custom, high-value EPIC, and EPIC).

| Locus | | # DMCs (# in INSPIRE that overlap with URECA) | eQTM (Number of DMCs) | | Location(s) | Primary Criteria | | | Functional Annotations | | | | | |
|---|---|---|---|---|---|---|---|---|---|---|---|---|---|---|
| | | | Nearest gene | pcHi-C target gene | | GWAS | EWAS | DMR | Open chromatin (ATAC-seq) | Enhancer (pcHi-C) | TFBS (ENCODE) | TSS (ROADMAP) | Poised enhancer | Active enhancer |
| *CISH* | URECA (Custom) | 11 | *CISH* (11) | *RASSF1* (2), *MANF* (4), *ZMYND10* (1), *HYAL1* (2) | Exons 2 and 3 | 0 | 0 | 11 | 0 | 11 | 10 | 0 | 11 | 2 |
| | INSPIRE (Custom) | 12 (9) | NA | NA | Exon 3 | 0 | 0 | 12 | 0 | 12 | 12 | 0 | 12 | 1 |
| | URECA (high-value EPIC) | 2 | *CISH* (2) | 0 | Exon 3, intron 1 | 0 | 2 [E24,E25] | 1 | 0 | 2 | 2 | 1 | 1 | 0 |
| | URECA (EPIC) | 1 | *CISH* (1) | *HYAL1* | Intergenic | 0 | 0 | 0 | 0 | 1 | 1 | 0 | 1 | 1 |
| *SLC22A5/ IRF1* | URECA (Custom) | 2 | 0 | *IRF1* (2), *CCNI2* (1), *GDF9* (2), *SEPT8* (1), *SHROOM1* (1) | Intron 6 | 2 [E9,E10] | 0 | 0 | 2 | 2 | 2 | 0 | 2 | 2 |
| | INSPIRE (Custom) | 1 (1) | NA | NA | Intron 6 | 1 [E9,E10] | 0 | 0 | 1 | 1 | 1 | 0 | 1 | 1 |
| | URECA (high-value EPIC) | 1 | 0 | *IRF1* (1), *CCNI2* (1) | Intron 6 | 1 [E9,E10] | 1 [E24,E25] | 0 | 1 | 1 | 1 | 0 | 1 | 1 |
| | URECA (EPIC) | 0 | 0 | 0 | - | | | | | | | | | |
| *HDAC7 /VDR* | URECA (Custom) | 3 | *HDAC7* (1) | *SLC48A* (1) | Intergenic | 3 [E9] | 0 | 3 | 0 | 3 | 3 | 0 | 0 | 3 |
| | INSPIRE (Custom) | 2 (2) | NA | NA | Intergenic | 2 [E9] | 0 | 2 | 0 | 2 | 2 | 0 | 0 | 2 |
| | URECA (high-value EPIC) | 1 | *VDR* (1) | 0 | Intergenic | 1 [E9] | 1[E25] | 0 | 0 | 1 | 1 | 0 | 0 | 1 |
| | URECA (EPIC) | 1 | *VDR* (1) | 0 | Intergenic | 1 [E9] | 0 | 0 | 0 | 1 | 1 | 0 | 0 | 1 |

**Supplementary Dataset 1 (separate file).** Information on 37,863 Custom Array CpGs that passed manufacture and array QC. Details regarding CpG inclusion based on primary criteria (EWAS, GWAS, or WGBS_DMR) and functional annotation categories are provided in columns G through Q. See **Methods** for detailed explanation of categories.

**Supplementary Dataset 2 (separate file).** List of 26,905 EPIC CpGs that meet criteria for inclusion on the Custom Array (i.e. Filtered EPIC). Details regarding primary criteria (EWAS, GWAS, or WGBS_DMR) and functional annotation categories are provided in columns G through Q. See **Supplementary Methods** for detailed explanation of categories.
